# Puberty Timing and Cognitive Functioning: Insights from the Adolescent Brain Cognitive Development (ABCD) Study and Mendelian Randomization

**DOI:** 10.1101/2025.07.10.25331286

**Authors:** Lars Dinkelbach, Bianca Serio, Sofie Valk, Franka Edith Weisner, Luise Bläschke, Triinu Peters, Anke Hinney, Börge Schmidt, Raphael Hirtz

**Affiliations:** Department of Pediatrics III, University Hospital Essen, University of Duisburg-Essen, Essen, Germany; Institute of Sex- and Gender-sensitive Medicine, University Hospital Essen, University of Duisburg-Essen, Essen, Germany; Institute of Neuroscience and Medicine, Brain & Behavior (INM-7), Research Centre Jülich, Jülich, Germany; Institute of Systems Neuroscience, Medical Faculty, Heinrich-Heine-Universität Düsseldorf, Düsseldorf, Germany; Max Planck School of Cognition, Leipzig, Germany; Max Planck Institute for Human Cognitive and Brain Sciences, Leipzig, Germany; Section of Molecular Genetics in Mental Disorders, University Hospital Essen, University of Duisburg-Essen, Essen, Germany; Center for Translational Neuro- and Behavioral Sciences, University Hospital Essen, University of Duisburg-Essen, Essen, Germany; Institute for Medical Informatics, Biometry and Epidemiology, University of Duisburg-Essen, Essen, Germany; Center for Child and Adolescent Medicine, Helios University Hospital Wuppertal, Witten/Herdecke University, Wuppertal, Germany

**Author notes:** **Corresponding Author:** Lars Dinkelbach, MD, BSc, Department of Pediatrics III, University Hospital Essen, University of Duisburg-Essen, Hufelandstr. 55, Essen, Germany.

**Keywords:** Adolescence, Puberty Timing, Cognition, Memory, Executive Functions

## Abstract

**Purpose:** Early puberty timing is associated with altered neurodevelopment and adverse mental health. Its association with cognitive functioning remains unclear. We investigated the effects of puberty timing on various cognitive abilities.

**Methods:** In 10,174 participants from the Adolescent Brain and Cognitive Development (ABCD) Study, we assessed associations between puberty timing at baseline (age 9.9±0.6 years) and performance on six cognitive tasks at baseline, 2-year, and 4-year follow-ups. Linear-mixed-regression models were calculated separately by sex, adjusting for bodyweight, birthweight, parental income, and race/ethnicity. To probe causal relationships, we performed Mendelian randomization utilizing external Genome Wide Association Study (GWAS) data on age at menarche (N=632,955), male puberty timing (N=205,354), and executive functioning (N=427,037).

**Results:** In the ABCD study, earlier puberty timing was associated with poorer performance pooled across timepoints on the NIH Toolbox® Picture Sequence Memory Task (girls: standardized β=−0.04, 95%-CI [−0.06, −0.01]; boys: β=−0.03, 95%-CI [−0.05, −0.01]), the List Sorting Working Memory Test (girls: β=−0.03, 95%-CI [−0.06, −0.01]; boys: β=−0.04, 95%-CI [−0.06, −0.01]), and both the learning of a word list (girls: β=−0.05, 95%-CI [−0.07, −0.02]; boys: β=−0.04, 95%-CI [−0.06, −0.01]) and its recall (girls: β=−0.03, 95%-CI [−0.06, −0.01]; boys: β=−0.03, 95%-CI [−0.06, −0.01]). Mendelian randomization indicated better executive functioning with later age at menarche (b=0.005/year, 95%-CI [0.000, 0.011]); the association with male puberty timing was directionally consistent but remained non-significant (b=0.012/year, 95%-CI [−0.004, 0.028]).

**Discussion:** These findings support a causal link between earlier puberty timing and poorer cognitive performance. However, effect sizes were small, indicating negligible clinical relevance.

## Background

Adolescence, initiated by puberty, is marked by profound physical, psychological, and social transitions that demand increasing cognitive flexibility. In this context, cognitive control processes or executive functions —such as response inhibition, working memory, and task switching—undergo rapid developmental gains during adolescence, likely reflecting concurrent neurobiological maturation [1–3].

Puberty timing—defined as the relative onset of puberty compared to same-aged, same-sex peers— varies substantially between individuals, shows sex differences, and has shifted toward earlier onset over time [4]. Early puberty in girls has consistently been associated with adverse mental health outcomes, including increased risk for depression, while findings for boys have been mixed [5–8].

Despite the known impact of puberty timing on mental health, its influence on cognitive function remains underexplored. Some studies have reported associations between early puberty and lower executive functioning [9] and academic performance [10], while others have found higher cognitive performance [11] and steeper gains in attention skills [12]. Some evidence suggests diverging sex-specific effects, with early puberty linked to poorer academic performance and self-control in girls [10, 12], and to better mental rotation skills in boys [13].

Puberty timing and cognitive function are closely intertwined with biological and environmental factors. Both early pubertal maturation and impaired cognitive function have been linked to increased weight [14, 15], low birthweight [16], and environmental influences including low socioeconomic status and social deprivation [17, 18]. These confounders are likely to contribute to mixed findings in the literature and further complicate causal inference.

Mendelian randomization (MR) studies take advantage of the random allocation of genetic variances during conception. If genetic variants are strongly linked to a phenotype of interest, e.g. puberty timing, those variants can serve as predictors for this phenotype, unconfounded by life experiences or environmental factors [19, 20]. Because genetic variants are fixed at conception, Mendelian randomization helps to overcome key limitations of observational research, particularly residual confounding and reverse causation, enabling causal inference [19, 20].

## Objective

Here, we aim to (1) investigate the association between puberty timing and specific cognitive functioning using large-scale longitudinal data from the ABCD Study, (2) establish causal relationships through comprehensive Mendelian randomization analyses, and (3) examine sex-specific patterns in these associations.

## Methods

### ABCD Study

#### Sample Description and Exclusion Criteria

The ABCD Study is a longitudinal cohort of 11,868 children and adolescents recruited from 21 sites across the United States, with repeated assessments focusing on brain development. We used data at baseline (up to N=10,174; age 9.9±0.6 years), 2-year follow-up (up to N=9,043; age 12.0±0.7 years), and 4-year follow-up (up to N=3,924; age 14.1±0.7 years). Details on inclusion and exclusion criteria are provided in Supplemental Methods 1.

#### Puberty Timing

Puberty timing at baseline was assessed using the parent report of the Pubertal Development Scale (PDS, [21]). We chose the parental version instead of the self-report because, as in late childhood/early adolescence, parental ratings show better agreement with physical changes measured by clinicians than self-report [22]. The PDS comprises five items with a total score ranging from 5 to 20 (Table 1). To derive a continuous measure of puberty timing relative to same-aged peers of the same sex, we first excluded subjects with missing puberty information at baseline (Supplementary Figure S1), then regressed the total PDS score at baseline on age separately by sex and used the standardized residuals for further analyses, following prior approaches [5]. Positive residuals indicate earlier puberty timing (Supplementary Figure S2 and S3).

**Table 1.**
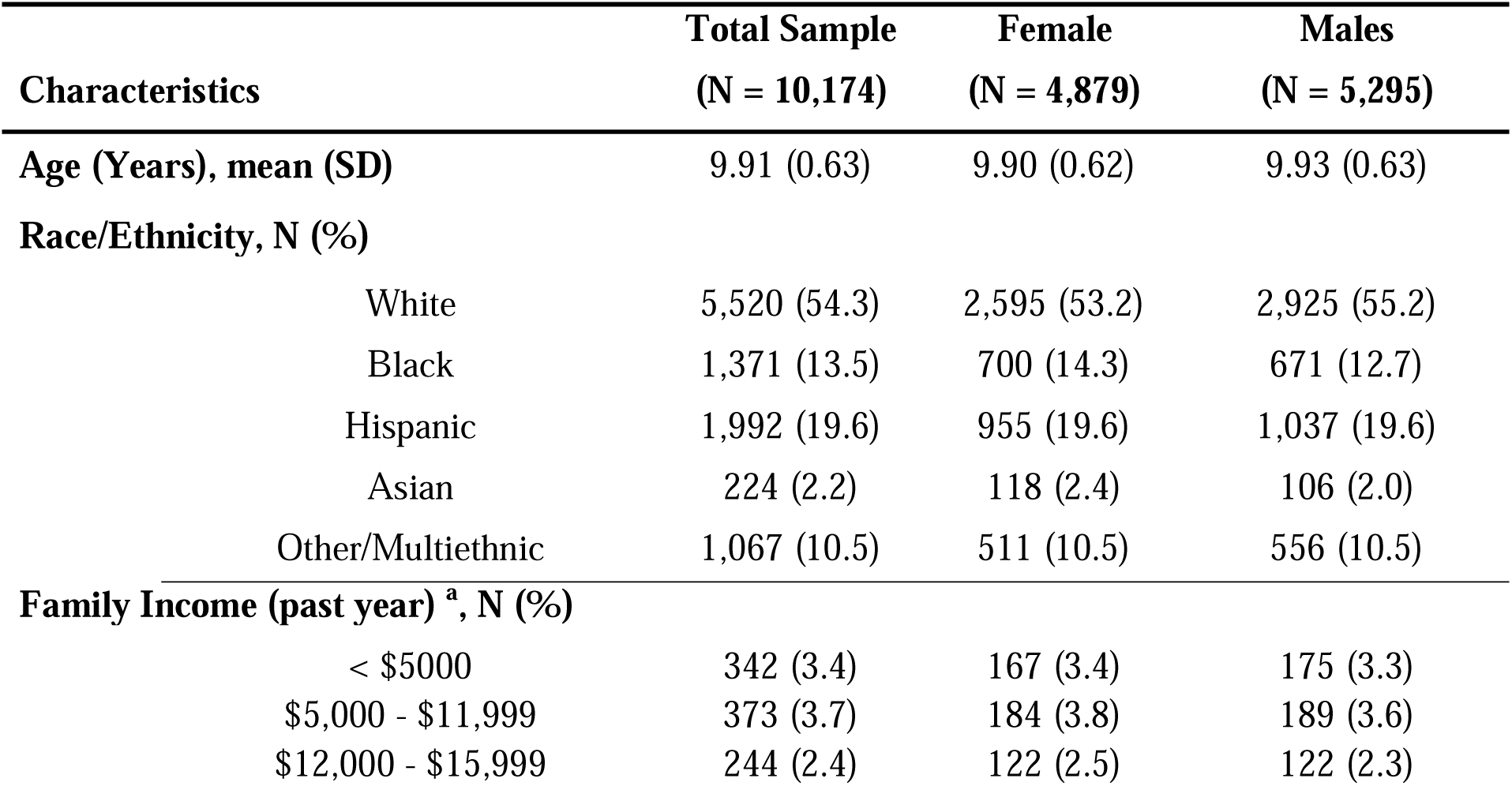
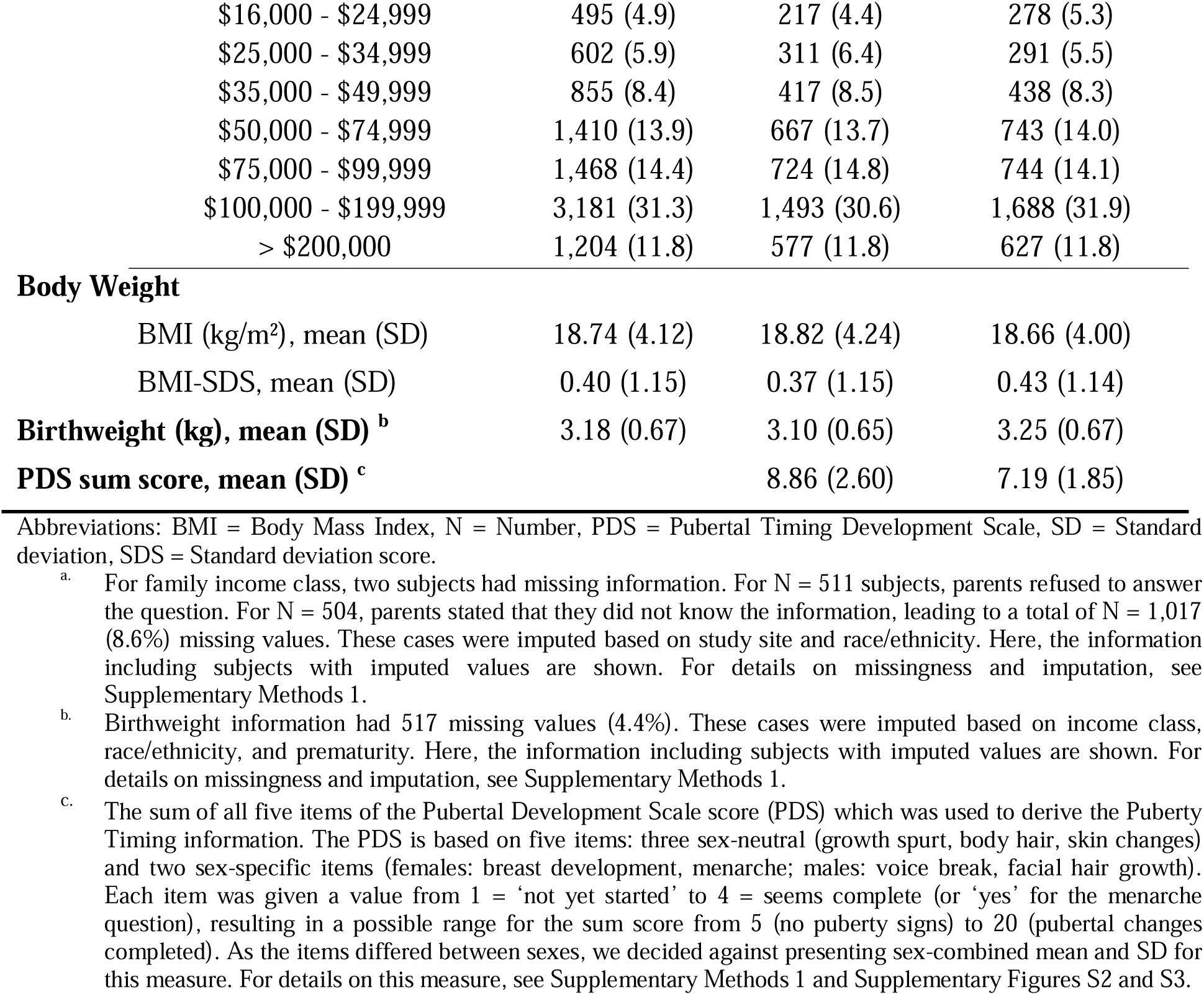
Baseline Characteristics of the final ABCD Sample.

#### Cognitive Measures

As cognitive outcome measures, the scores on the following tasks were used: from the NIH Toolbox® [23, 24], the Flanker Task, Pattern Comparison Task, the Picture Sequence Memory Task, and the List Sorting Task; the Rey Auditory Verbal Learning Test (RAVLT); and the Little Man Task (LMT [25, 26]). Figure 1 illustrates the tasks and their instructions; detailed task characteristics are reported elsewhere [2, 27]. While the Flanker and List Sorting Tasks assess core components of executive functioning—cognitive control and working memory—the remaining tasks primarily target episodic memory (Picture Sequence Memory and RAVLT), processing speed (Pattern Comparison), and visuospatial processing (LMT).

**Figure 1.**
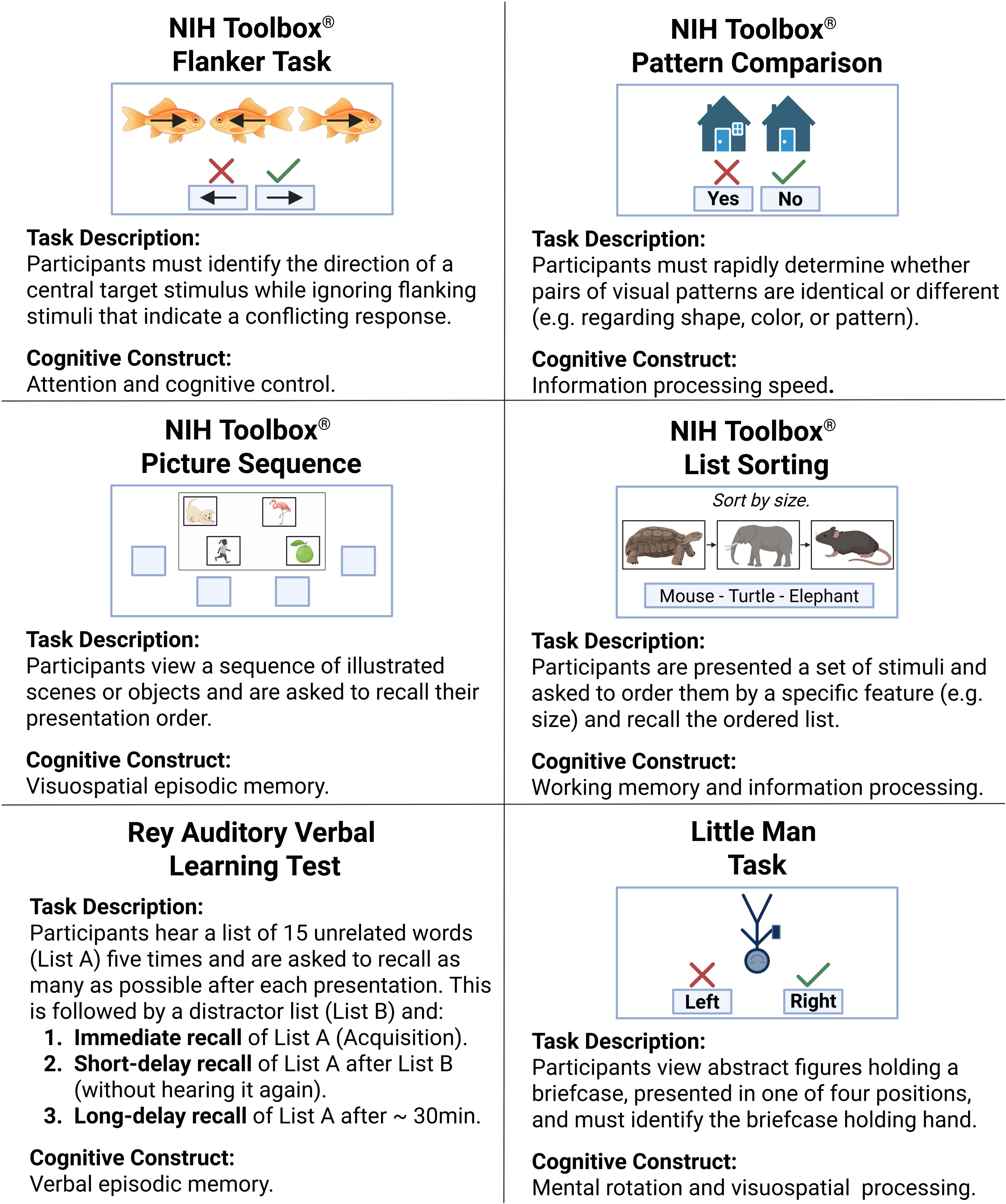
Description of Cognitive Tasks. This figure depicts the cognitive tasks used in the Adolescent Brain Cognitive Development (ABCD) Study. A more detailed description of the tasks can be found in Supplementary Methods 1. NIH Toolbox® Flanker Task = NIH Toolbox® Flanker Inhibitory Control and Attention Test; NIH Toolbox® Pattern Comparison = NIH Toolbox® Pattern Comparison Processing Speed Test; NIH Toolbox® Picture Sequence = NIH Toolbox® Picture Sequence Memory Test; NIH Toolbox® List Sorting = NIH Toolbox® List Sorting Working Memory Test. The NIH Toolbox® tasks are a trademark of © 2025 Toolbox Assessments, Inc. All rights reserved. The images serve only as illustrative examples and do not represent the actual content of the tasks.

#### Covariates and Statistical Analysis

Given the varying pubertal development trajectories between sexes, all analyses were run separately by sex. After confirming the normal distribution of residuals and testing for multicollinearity (Supplementary Table S5, Supplementary Figures S9 and S10), multiple (for each cognitive outcome) univariate mixed linear models were run to assess the effect of baseline puberty timing as a time-invariant fixed effect on each of nine cognitive outcome measures, both overall (i.e., across all time points), and separately by event (baseline, 2-year follow-up, and 4-year follow-up). For each outcome, linear-mixed regression models were run, which included fixed effects for baseline puberty timing, the BMI-Standard Deviation Scores (BMI-SDS), birthweight, combined family income, and race/ethnicity (5-category factor), and a random intercept for family nested in site (and, for the overall analysis, additional random intercepts for subject and timepoint). This adjustment set was derived by constructing a directed acyclic graph (DAG) to identify the minimal sufficient set for estimating the unconfounded total effect of puberty timing on cognitive functioning (see Supplementary Figure S7 for details and assumptions). Given the relatively high number of missing values for birthweight (N=517; 4.4%) and family income (N=1,017; 8.6%), missing data for these two variables were imputed (Supplementary Methods 1).

#### Sensitivity Analyses

To assess whether mental health adversity influenced the relationship between puberty timing and cognitive task performance, we extended our models by including the externalizing and internalizing symptom scores from the parent-reported of the Child Behavior Checklist (CBCL) as fixed effects. To examine potential non-linearity in these associations, i.) Generalized Additive Mixed Models (GAMMs) were fitted that allowed for a smoothed term for puberty timing, and ii.) raw cognitive task scores were plotted against puberty timing to detect any non-linear trends. To assess whether the limited variability in male puberty timing at baseline (age 9.9±0.6 years) affected results, the analyses in males were re-ran using puberty timing measured at the 2-year follow-up (age 12.0±0.7 years).

#### Ethics

Written informed consent was obtained from legal guardians and assent from all participating children. The ABCD protocol was approved by the central IRB at UC San Diego and by local IRBs at some sites. Data use for this study was approved by the Ethics Committee of the University of Duisburg-Essen (24-12040-BO).

### Mendelian Randomization Analyses

#### MR Assumptions

MR uses genetic variants—or Single Nucleotide Polymorphisms (SNPs)—as instrumental variables to estimate the effect of an exposure (puberty timing) on an outcome (executive functioning). Because these variants are randomly allocated at conception and are independent of subsequent environmental factors, they can serve as unbiased proxies for puberty timing, reducing residual confounding and enabling causal inferences. Valid SNP-based instrumental variables must satisfy three assumptions: i.) *relevance*, meaning the genetic variants are strongly associated with the exposure (defined by F-statistics > 10 [19]); ii.) *independence*, indicating no association with confounders of the exposure–outcome relationship; and iii.) *exclusion restriction*, postulating that the genetic variants influence the outcome only through the exposure [19, 20].

#### Exposure: Puberty Timing

For females, data of a GWAS on age at menarche from up to 632,955 participants of European ancestry from the UK Biobank, 23andMe, Inc., and additional cohorts was used [28], with 971 independent genome-wide significant (p<5×10^−8^) SNPs for age at menarche of which 832 SNPs (85.7%) were available for analysis (Supplementary Methods 2).

For males, a GWAS on 205,354 men of European ancestry on male puberty timing was used, combining data from the UK Biobank and 23andMe, Inc. Male puberty timing was assessed via recalled age at voice break (23andMe) or self-reported early/late/average onset of voice break and facial hair growth (UK Biobank), and these phenotypes were meta-analyzed to derive a composite measure, leading to the identification of 76 genome-wide significant independent lead SNPs (72 (94.7%) available for analysis) [29].

#### Outcome: Executive Functioning

To capture executive functioning, a GWAS on 427,037 participants of the UK Biobank was used [30]. Here, a phenotypic factor score for executive functioning was derived using a confirmatory factor analysis based on five neuropsychological tasks (for a description of the tasks, see [30]).

Although based on a different set of tasks, the operationalization of executive functioning in the GWAS showed considerable conceptual overlap with the cognitive domains assessed in the ABCD study, particularly regarding cognitive control, processing speed, and working memory.

#### Sensitivity Analyses

The Inverse Variance Weighted (IVW) method was employed as primary analysis. To face potential violations of the Mendelian randomization assumptions, which often arise from pleiotropy (where one genetic variant affects multiple traits), we took the following steps: i.) pleiotropic SNPs identified by MR-PRESSO were excluded; ii.) pleiotropy-robust methods—MR-Egger, Weighted Median, Penalized Weighted Median, and Lasso Penalization—were conducted as sensitivity analyses; iii.) given the strong genetic overlap between puberty timing and body weight [28, 29] we additionally adjusted for BMI in multivariable Mendelian randomization [31].

Because all three GWASs—age at menarche, male puberty timing, and executive functioning— included UK Biobank participants, sample overlap may have introduced bias [32]. Therefore, we used MR-APSS, which accounts for both, bias from sample overlap and pleiotropy [33]. However, MR-APSS requires full summary statistics for the exposure, which were available only for age at menarche. To mitigate sample overlap in males, we performed Mendelian randomization analyses using only age at voice break data from 23andMe (N=55,871 [29]). Of the 76 male puberty timing SNPs, 36 (F-statistics > 10) were valid instruments for age at voice break.

To ensure that the exposure influences the outcome and not vice-versa, we conducted reverse Mendelian randomization using executive functioning as the exposure and age at menarche as the outcome. This analysis relies on the availability of full summary statistics and thus was conducted for age at menarche only. For further details on all methods, see Supplementary Methods 2.

## Results

### ABCD sample analyses

Linear mixed models assessing the effect of puberty timing across all time points (i.e., the ‘Overall’ model) revealed a decline in task performance with earlier puberty timing in girls and boys with regard to: the NIH Toolbox® Picture Sequence Task (girls: standardized beta, i.e. SD change of the outcome per SD change of the exposure, β=−0.04, 95%-CI [−0.06, −0.01]; boys: β=−0.03, 95%-CI [−0.05, −0.01], the NIH Toolbox® List Sorting Test (girls: β=−0.03, 95%-CI [−0.06, −0.01]; boys: β=−0.04, 95%-CI [−0.06, −0.01]), and the number of recalled items in the RAVLT after learning trials (RAVLT – Acquistion, girls: β=−0.05, 95%-CI [−0.07, −0.02]; boys: β=−0.04, 95%-CI [−0.06, −0.01]) and after a ∼30min delay (RAVLT – Delayed Recall, girls: β=−0.03, 95%-CI [−0.06, −0.01]; boys: β=−0.03, 95%-CI [−0.06, −0.01]), see Figure 2 and 3 and Supplementary Table S8. In girls, earlier puberty timing was also associated with reduced accuracy (β=−0.03, 95%-CI [−0.05, −0.01]) but shortened response time (β=−0.03, 95%-CI [−0.05, −0.01]) in the mental rotation task (LMT). After controlling response time in an additional linear mixed model, the negative effect of early puberty timing on LMT-Accuracy remained (β=−0.03, 95%-CI [−0.05, −0.00]).

**Figure 2.**
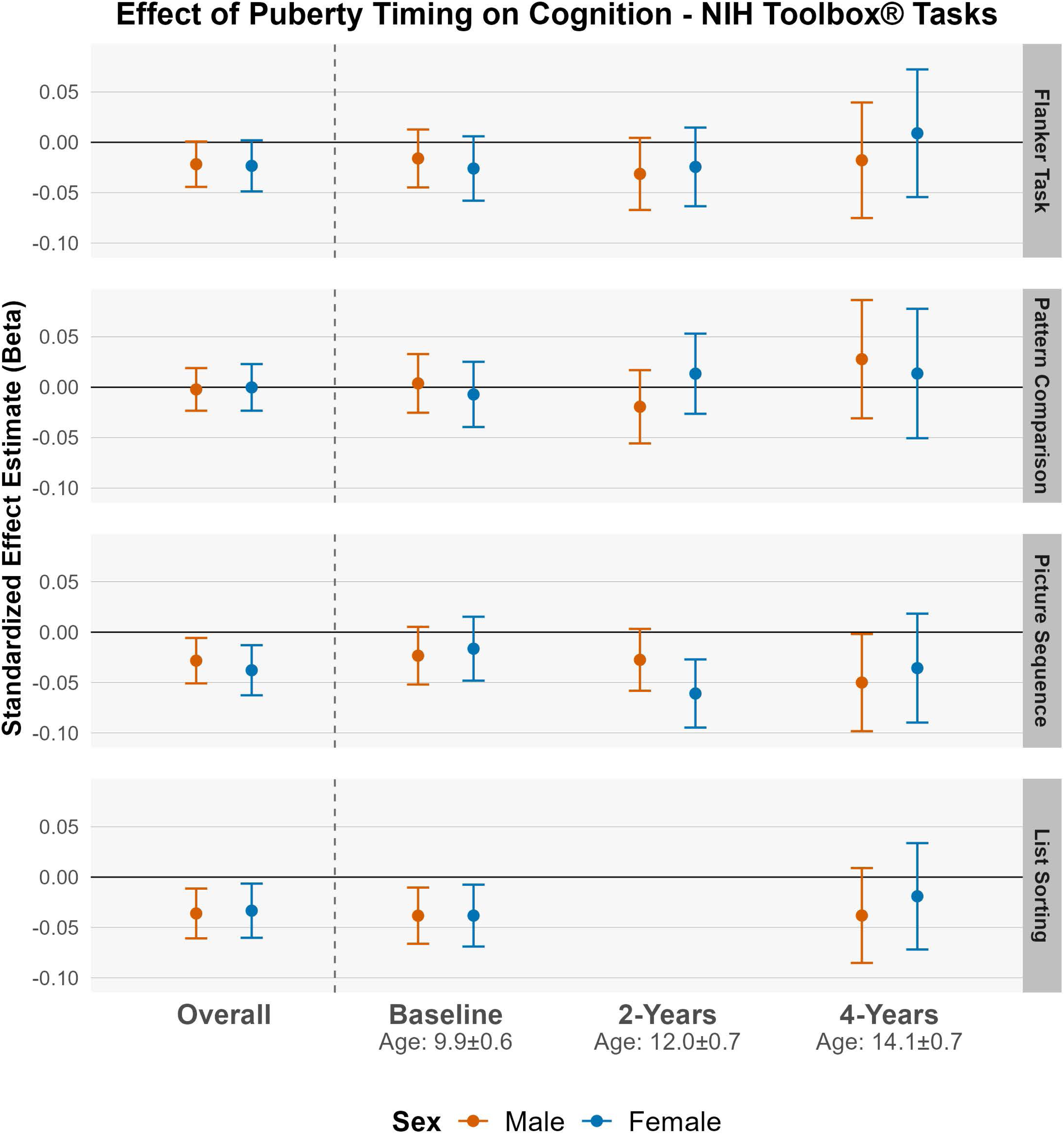
Effect of Puberty Timing on Cognition - NIH Toolbox® Tasks. This figure illustrates the association between puberty timing—quantified as standardized residuals from the regression of the Pubertal Development Scale (PDS) item sum on age, reflecting physical pubertal maturation relative to same-age, same-sex peers—and uncorrected standard scores from cognitive tasks in the NIH Toolbox®. Displayed are standardized effect sizes (i.e., the standard deviation [SD] change in cognitive performance per SD increase in Puberty Timing), derived from linear mixed models. These models were adjusted for BMI-Standard Deviations Scores, race/ethnicity, family income class, and birthweight, and included a random intercept for family nested within study site. The ‘Overall’ model also included random intercepts for subject (to account for repeated measures) and event (to capture potential task-specific variation, such as minor implementation changes; see [2]). Flanker Task = NIH Toolbox^®^ Flanker Inhibitory Control and Attention Test; Pattern Comparison = NIH Toolbox^®^ Pattern Comparison Processing Speed Test; Picture Sequence = NIH Toolbox^®^ Picture Sequence Memory Test; List Sorting = NIH Toolbox^®^ List Sorting Working Memory Test.

**Figure 3.**
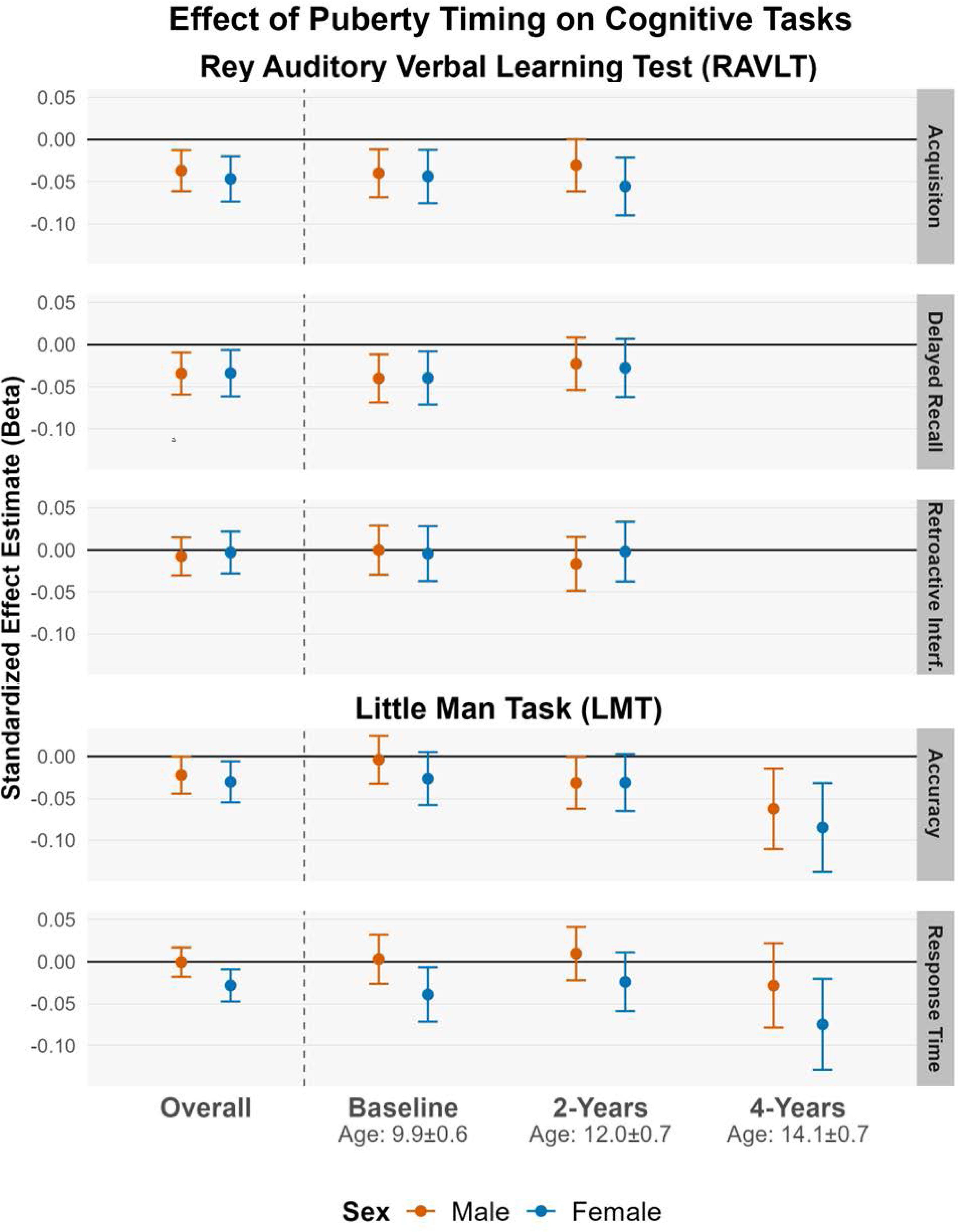
Effect of Puberty Timing on Cognition – RAVLT and LMT. This figure depicts the effect of puberty timing on performance in the Rey Auditory Verbal Learning Test (RAVLT) and the Little Man Task (LMT). Shown are standardized effect sizes, representing the standard deviation change in cognitive task performance per standard deviation increase in Puberty Timing, derived from adjusted linear mixed models. In the RAVLT, Acquisition refers to the total number of correctly recalled items from List A after five learning trials, reflecting initial learning ability. Delayed Recall represents the number of correctly recalled items from List A following a delay of approximately 30 minutes, serving as a measure of episodic memory. Retroactive Interference is calculated as the difference between the number of correctly recalled items from List A after five learning trials and the number recalled after the presentation of a distractor list. Higher values indicate greater interference and therefore poorer memory performance. In the LMT, the Accuracy score corresponds to the number of correct responses in the mental rotation task, while the Response Time refers to the average time taken to produce correct responses.

#### Effect of Puberty Timing on Cognitive Functioning at consecutive Timepoints

The timepoint-specific analyses revealed mixed findings: for the NIH Toolbox® Picture Sequence Task, effect sizes remained stable across timepoints (girls β from −0.02 to −0.06; boys β from −0.02 to −0.05, Figure 2-3 and Supplementary Table S9), with significant associations at the 2-year follow-up in girls (β=−0.06, 95%-CI [−0.09, −0.03]) and at the 4-years follow-up in boys (β=−0.05, 95%-CI [−0.10, −0.00]). Similarly, puberty timing was negatively associated with RAVLT-Acquisition at baseline (girls: β=−0.04, 95%-CI [−0.08, −0.01], boys: β=−0.04, 95%-CI [−0.07, −0.01]) and at the 2-year follow-up in girls (β=−0.06, 95%-CI [−0.09, −0.02]), with borderline result in boys (β=−0.03, 95%-CI [−0.06, 0.00]). For the NIH Toolbox® List Sorting Test (girls and boys: β=−0.04, 95%-CI [−0.07, −0.01]) and for the RAVLT-Delayed Recall performance (girls and boys: β=−0.04, 95%-CI [−0.07, −0.01]), negative effects were observed only at baseline, but not at subsequent follow-ups. In the mental rotation task (LMT), puberty timing was negatively associated with accuracy at the 4-year follow-up (girls: β=−0.08, 95%-CI [−0.14, −0.03], boys: β=−0.06, 95%-CI [−0.11, −0.01]), whereas effects at baseline and 2-year follow-up were smaller (girls: baseline β=−0.03, 95%-CI [−0.06, 0.01]; 2-year β=−0.03, 95%-CI [−0.06, 0.00]; boys: baseline β=−0.00, 95%-CI [−0.03, 0.02]; 2-year β=−0.03, 95%-CI [−0.06, −0.00].

#### Sensitivity Analysis

To examine whether effects were driven by mental health, we added CBCL internalizing and externalizing scores to the linear mixed models, which had no substantial impact on effect estimates (Supplementary Table S8). To assess non-linearity, we ran GAMMs and inspected plots of cognitive scores against puberty timing; model fit of non-linear models showed no consistent improvement (Supplementary Table S6), and visual patterns supported linear associations (Supplementary Figures S11–S12). Using puberty timing at the 2-year follow-up in males yielded similar results, with earlier timing linked to lower performance in the NIH Toolbox® Picture Sequence Task, the NIH Toolbox® List Sorting Task, and RAVLT-Acquisition (Supplementary Figure S13 and Supplementary Table S7).

### Results of Mendelian Randomization analyses

For age at menarche, the IVW estimate revealed a significant positive effect on executive functioning, indicating that early menarche is associated with poorer executive functioning (b=0.005 per-one-year increase in age at menarche, 95%-CI [0.000, 0.011], Figure 4). The Q-statistic indicated significant heterogeneity (Q-statistic=1943.2, p=6.9×10^-96^). All conducted sensitivity analyses lead to similar findings for the effect of age at menarche. Reverse Mendelian randomization, using executive functioning as the exposure and age at menarche as the outcome, did not provide consistent evidence for reverse causality (b=−0.02, 95%-CI [−0.17, 0.13], Supplementary Figure S17).

**Figure 4.**
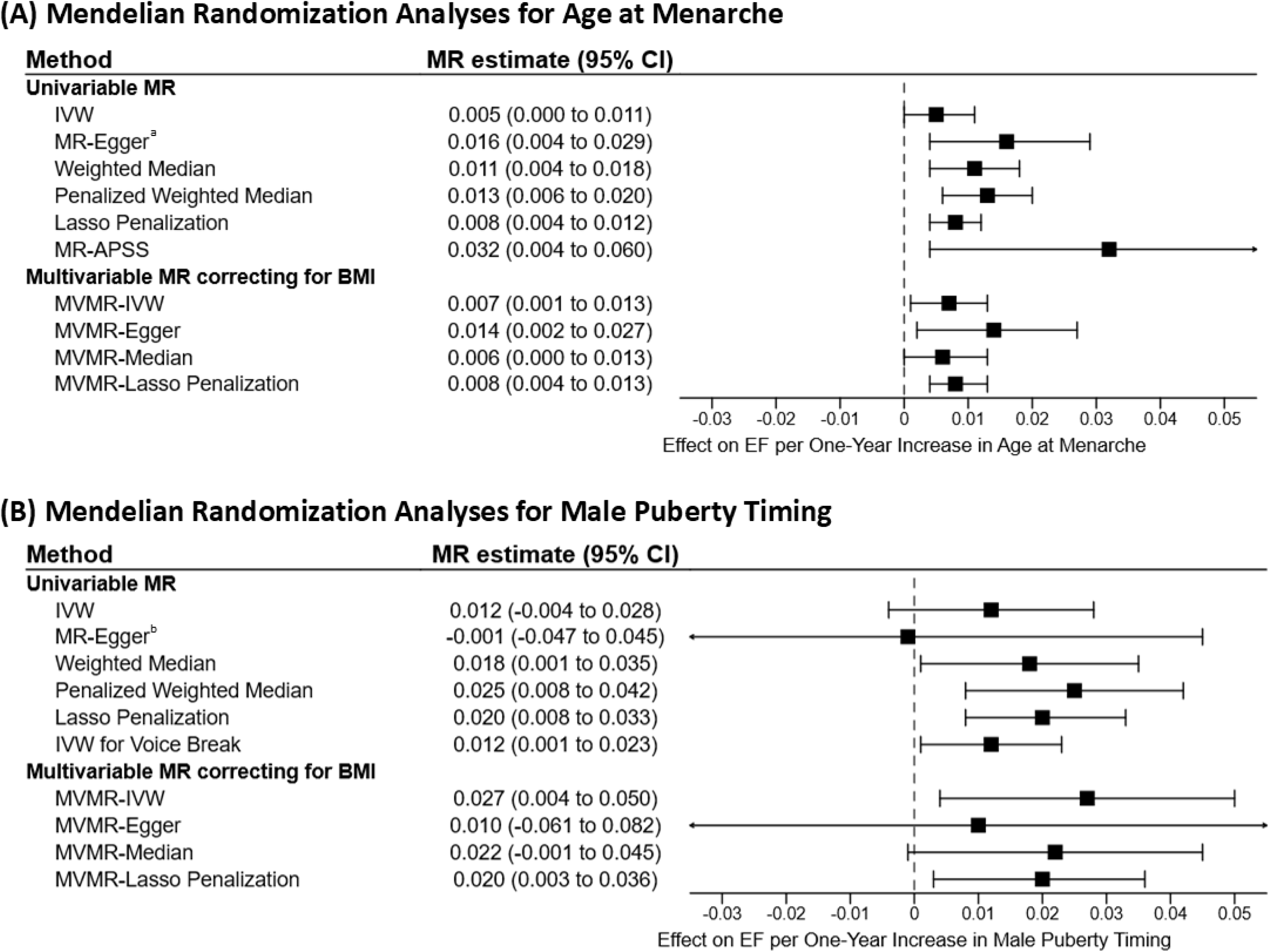
Results of Mendelian Randomization (MR) analyses. This figure illustrates the effect of female puberty timing (measured by the age at menarche) and male puberty timing on executive functioning (EF). The MR effect estimate (b) represents the effect estimate of one year increase in Age at Menarche/Male Puberty Timing on EF. *a:* Eggers-intercept did not show evidence for significant directional pleiotropy (intercept=−0.0003, se=0.0002, p=0.063). The *I*^2^-statistic was 0.900, indicating a near-threshold violation of the ‘no measurement error’ (NOME) assumption. *b:* Eggers-intercept did not show evidence for significant directional pleiotropy (intercept= 0.0004, se=0.007, p=0.564). The *I*^2^-statistic was 0.741, indicating a violation of the NOME assumption. Therefore, the MR Egger result should be interpreted with caution. For explanation of the different Mendelian Randomization methods used, see Supplementary Methods 2.

The effect estimate for male puberty timing on executive functioning was slightly larger but had wider confidence intervals, resulting in a non-significant finding (IVW, Effect on executive functioning per-one-year increase in male puberty timing b=0.012, 95%-CI [−0.004, 0.028]). The Q-statistic indicated significant heterogeneity (Q-statistic=153.9, p=3.1×10^-8^). Sensitivity analyses yielded mixed results; MR-Egger showed a near-zero effect with wide confidence intervals. However, all other sensitivity analyses, including heterogeneity-robust methods, indicated an effect of male puberty timing on executive functioning (b ranging from 0.012 to 0.027, Figure 4).

## Discussion

Previous studies on puberty timing and cognitive performance yielded conflicting results. In this study, we combined comprehensive observational data from the ABCD Study with Mendelian randomization analyses to assess whether puberty timing is associated with cognitive performance.

### Early Puberty Timing leads to negative Cognitive Performance

In this study of 10,174 participants from the ABCD Study, and after adjustment for relevant confounders, we observed a negative association between puberty timing and performance, whereby earlier puberty timing was systematically related to lower cognitive functioning, consistent with previous findings linking early maturation to poorer academic performance and executive functioning [9, 10]. The negative effects were most pronounced in tasks assessing episodic and working memory, with earlier puberty timing associated with poorer performance on the NIH Toolbox® Picture Sequence Memory Task (visuospatial episodic memory), List Sorting Working Memory Test (working memory and information processing), and both learning and delayed recall in the RAVLT (verbal episodic memory) in both sexes. Apart from faster response times in the mental rotation task (LMT), which were accompanied by reduced accuracy, there was no support for positive effects of early puberty timing on cognitive abilities, contrasting earlier reports of positive associations [11, 13]. These observational findings were supported by Mendelian randomization analyses, which showed that earlier age at menarche was associated with lower executive functioning, an effect robust across multiple sensitivity analyses accounting for pleiotropy and sample overlap. Furthermore, reverse Mendelian randomization did not support a causal effect in the opposite direction (i.e., lower executive functioning leading to earlier age at menarche). Together, these results provide evidence for a causal, directional of earlier puberty timing leading to lower cognitive performance, at least in some cognitive domains.

### Puberty Timing and Cognition: Limited Evidence for Sex-Specific Effects

Several studies have suggested sex-specific effects of puberty timing on cognitive abilities [10, 12], such as poorer self-control in early-maturing girls [12] and better mental rotation skills in early-maturing boys [13]. Here, Mendelian randomization analyses showed an effect of age at menarche on executive functioning but provided inconsistent results for males, this should not be interpreted as a sex-specific effect. First, the discrepant finding may reflect differences in power due to sample size (age at menarche: N=632,955; male puberty timing: N=205,354). Second, sensitivity analyses robust to pleiotropy indicated a negative effect of earlier male puberty timing on executive functioning, consistent with the findings in females. Third, earlier puberty timing was associated with poorer cognitive performance across multiple tasks with comparable effect sizes in both sexes in the ABCD Study. Overall, these findings do not support systematic sex-specific effects but rather suggest that earlier puberty timing adversely affects cognitive functioning in both sexes.

### Mixed Evidence for Long-Term Effects of Puberty Timing on Cognitive Abilities

In the ABCD Study, timepoint-specific results showed mixed findings regarding the stability of the association between puberty timing and cognitive performance: for some tasks (e.g., NIH Toolbox® List Sorting), the effect of puberty timing attenuated over time, while for others, it remained stable (e.g., NIH Toolbox® Picture Sequence Task) or even increased (e.g., LMT-Accuracy) over the 4-year study period. The latter finding is consistent with the Mendelian randomization analysis, suggesting that effects on executive functioning may persist into late adulthood at the age of ∼40–69.

### Small Effect Sizes - Limited Clinical Significance

The increasing availability of large-scale data demands careful interpretation of statistical findings that meet conventional significance thresholds. This study indicates that earlier puberty timing is associated with reduced performance across a range of cognitive tasks, based on two independent approaches (observational data and Mendelian randomization). However, effect sizes were small, with beta coefficients (i.e., SD change in cognitive performance per SD earlier puberty timing) ranging from −0.03 to −0.05 for statistically significant findings (in the ABCD study, after covariate adjustment), suggesting negligible functional relevance of puberty timing within normal ranges. However, our findings indicate a principal mechanism of altered cognitive functioning with puberty timing, which might be relevant under specific conditions, e.g., in cases of markedly early puberty, such as in central precocious puberty, and highlight the need to study cognitive effects of puberty suppression with GnRH analogues [34].

### Potential Role of Frontal Neurodevelopment in Early Puberty-Associated Cognitive Decline

Neurodevelopmentally, puberty is characterized by cortical thinning, increased myelination, synaptic pruning, and reorganization of functional neuronal networks, with the prefrontal cortex among the last regions to mature [3]. Executive functioning and working memory are strongly linked to prefrontal cortex development [35]. Adolescence has been proposed as a sensitive period for cognitive maturation, particularly for episodic and working memory formation [36]. Pubertal hormones may prematurely close this window of plasticity in early maturers [37]. Consistent with this, early puberty timing has been associated with altered brain morphology, notably greater cortical thinning in frontal regions [38], and reduced connectivity between frontal-cingulate and subcortical structures such as the hippocampus [39]. Further research is needed to determine whether these neurodevelopmental changes contribute to the subtle cognitive deficits associated with early puberty timing observed in the present study.

### Limitations

The present study has several limitations. First, it relies on indirect measures of puberty timing. In the ABCD Study, pubertal development was assessed through parent-reported physical maturation, while the Mendelian randomization analyses used recalled age at menarche (for females) and voice break or facial hair growth (for males). Thus, neither approach employed the gold standard of Tanner staging by a clinician to measure puberty timing.

Second, in the ABCD study, puberty timing was assessed at approximately 10 years of age, which may be too early to fully capture gonadal maturation in boys. At this stage, PDS scores may primarily reflect adrenarcheal changes, which typically begin around ages 6–8 [40]. However, this early assessment allowed us to examine longitudinal associations with cognitive outcomes. Importantly, results remained consistent in a sensitivity analysis using puberty timing measured at the 2-year follow-up in males (Supplementary Table S7).

Third, although both approaches—the observational ABCD Study and MR—led to similar conclusions, namely that earlier puberty timing is associated with impaired performance in some cognitive domains with small effect sizes, differences between the utilized measures should be noted. The executive functioning factor score used in the Mendelian randomization outcome GWAS [30] was derived from performance across five distinct cognitive tests and was assessed in middle aged adults (∼40–69 years, UK Biobank). While cognitive domains assessed in the UK Biobank largely overlap with those assessed in children and adolescents in the ABCD Study, they are not directly comparable, particularly due to the factor-analytic method used to construct the composite score of executive functioning [30].

## Conclusions

In this study of 10,174 participants from the ABCD Study, earlier puberty timing was associated with poorer cognitive performance, particularly in tasks related to episodic or working memory. Mendelian randomization analyses suggest, at least in females, a causal and directional association between puberty timing and executive functioning, with earlier age at menarche leading to decreased executive functioning in late adulthood. However, effect sizes with both approaches were small. Thus, while this evidence supports a causal influence of puberty timing on cognitive performance, the effect is likely negligible for everyday functioning for puberty timing within normal ranges. Further research should investigate underlying mechanisms and effects in pathologically altered puberty, e.g. in individuals with precocious puberty.

## Data Availability

Eligible researchers may apply for data access at the NIMH Data Archive (NDA) for the ABCD data used in this study (release 5.1, 10.15154/z563-zd24). The GWAS data used in this study was publicly available (see Supplementary Methods 2 for details). The code for statistical analyses may be shared upon reasonable request.

## Abbreviations

95%-CI: 95% confidence interval
ABCD Study: Adolescent Brain and Cognitive Development Study
BMI: Body Mass Index
EF: Executive Functioning
GWAS: Genome Wide Association Study
IVW: Inverse Variance Weighted
LMT: Little Man Task
MVMR: multivariable Mendelian randomization
N: Number
NIH: National Institutes of Health
PDS: Pubertal Timing Development Scale
RAVLT: Rey Auditory Verbal Learning Test
SD: Standard Deviation
SDS: Standard Deviation Score
SNP: Single Nucleotide Polymorphism

## Author Contributions

RH, and LD conceptualized the study. LD designed statistical analysis and conducted statistical analyses. LD wrote the first draft of the manuscript. BS, SV, FEW, LB, TP, AH, BS, and RH critically revised the manuscript.

## Acknowledgments

Data used in the preparation of this article were obtained from the Adolescent Brain Cognitive DevelopmentSM (ABCD) Study (Protected link to abcdstudy.org), held in the NIMH Data Archive (NDA). This is a multisite, longitudinal study designed to recruit more than 10,000 children age 9-10 and follow them over 10 years into early adulthood. The ABCD Study® is supported by the National Institutes of Health and additional federal partners under award numbers U01DA041048, U01DA050989, U01DA051016, U01DA041022, U01DA051018, U01DA051037, U01DA050987, U01DA041174, U01DA041106, U01DA041117, U01DA041028, U01DA041134, U01DA050988, U01DA051039, U01DA041156, U01DA041025, U01DA041120, U01DA051038, U01DA041148, U01DA041093, U01DA041089, U24DA041123, U24DA041147. A full list of supporters is available at https://abcdstudy.org/about/federal-partners/. A listing of participating sites and a complete listing of the study investigators can be found at https://abcdstudy.org/wp-content/uploads/2019/04/Consortium_Members.pdf. ABCD consortium investigators designed and implemented the study and/or provided data but did not necessarily participate in the analysis or writing of this report. This manuscript reflects the views of the authors and may not reflect the opinions or views of the NIH or ABCD consortium investigators. The ABCD data repository grows and changes over time. The ABCD data used in this report came from release 5.1, NIMH Data Archive Digital Object Identifier <u>10.15154/z563-zd24</u>.

This work was supported by a fellowship of the University Medicine Essen clinician scientist Academy (UMEA) to LD which was supported by the German Research Foundation (Deutsche Forschungsgemeinschaft; FU 356/12-2). The manuscript’s language style was enhanced through the utilization of artificial intelligence (ChatGTP). Figures were created with BioRender.com.

## References

1. Tervo-Clemmens B, Calabro FJ, Parr AC, Fedor J, Foran W, Luna B (2023) A canonical trajectory of executive function maturation from adolescence to adulthood. Nature communications 14:6922

2. Anokhin AP, Luciana M, Banich M, Barch D, Bjork JM, Gonzalez MR, et al. (2022) Age-related changes and longitudinal stability of individual differences in ABCD Neurocognition measures. Developmental Cognitive Neuroscience 54:101078

3. Arain M, Haque M, Johal L, Mathur P, Nel W, Rais A, et al. (2013) Maturation of the adolescent brain. Neuropsychiatric disease and treatment:449–461

4. Eckert-Lind C, Busch AS, Petersen JH, Biro FM, Butler G, Bräuner EV, et al. (2020) Worldwide secular trends in age at pubertal onset assessed by breast development among girls: a systematic review and meta-analysis. JAMA pediatrics 174:e195881–e195881

5. Ullsperger JM, Nikolas MA (2017) A meta-analytic review of the association between pubertal timing and psychopathology in adolescence: Are there sex differences in risk? Psychological bulletin 143:903

6. Dinkelbach L, Peters T, Grasemann C, Hinney A, Hirtz R (2025) The causal role of male pubertal timing for the development of externalizing and internalizing traits: results from Mendelian randomization studies. Psychological Medicine 55:e101

7. Hirtz R, Hars C, Naaresh R, Laabs B-H, Antel J, Grasemann C, et al. (2022) Causal Effect of Age at Menarche on the Risk for Depression: Results From a Two-Sample Multivariable Mendelian Randomization Study. Frontiers in Genetics 13

8. Hirtz R, Libuda L, Hinney A, Föcker M, Bühlmeier J, Holterhus P-M, et al. (2022) Age at menarche relates to depression in adolescent girls: Comparing a clinical sample to the general pediatric population. Journal of Affective Disorders 318:103–112

9. Porter BM, Roe MA, Mitchell ME, Church JA (2024) A longitudinal examination of executive function abilities, attention[deficit/hyperactivity disorder, and puberty in adolescence. Child Development 95:1076–1091

10. Suutela M, Miettinen PJ, Kosola S, Rahkonen O, Varimo T, Tarkkanen A, et al. (2022) Timing of puberty and school performance: A population-based study. Frontiers in Endocrinology 13:936005

11. Koerselman K, Pekkarinen T (2018) Cognitive consequences of the timing of puberty. Labour Economics 54:1–13

12. Chaku N, Hoyt LT (2019) Developmental trajectories of executive functioning and puberty in boys and girls. Journal of youth and adolescence 48:1365–1378

13. Beltz AM, Berenbaum SA (2013) Cognitive effects of variations in pubertal timing: is puberty a period of brain organization for human sex-typed cognition? Hormones and behavior 63:823–828

14. Reinehr T, Roth CL (2019) Is there a causal relationship between obesity and puberty? The Lancet Child & Adolescent Health 3:44–54

15. Likhitweerawong N, Louthrenoo O, Boonchooduang N, Tangwijitsakul H, Srisurapanont M (2022) Bidirectional prediction between weight status and executive function in children and adolescents: a systematic review and meta[analysis of longitudinal studies. Obesity Reviews 23:e13458

16. Miller S, DeBoer M, Scharf R (2018) Executive functioning in low birth weight children entering kindergarten. Journal of perinatology 38:98–103

17. Senger-Carpenter T, Seng J, Herrenkohl TI, Marriott D, Chen B, Voepel-Lewis T (2024) Applying life history theory to understand earlier onset of puberty: An adolescent brain cognitive development cohort analysis. Journal of Adolescent Health 74:682–688

18. Lawson GM, Hook CJ, Farah MJ (2018) A meta[analysis of the relationship between socioeconomic status and executive function performance among children. Developmental science 21:e12529

19. Lawlor DA, Harbord RM, Sterne JA, Timpson N, Davey Smith G (2008) Mendelian randomization: using genes as instruments for making causal inferences in epidemiology. Statistics in medicine 27:1133–1163

20. Haycock PC, Burgess S, Wade KH, Bowden J, Relton C, Davey Smith G (2016) Best (but oft-forgotten) practices: the design, analysis, and interpretation of Mendelian randomization studies. The American journal of clinical nutrition 103:965–978

21. Petersen AC, Crockett L, Richards M, Boxer A (1988) A self-report measure of pubertal status: Reliability, validity, and initial norms. Journal of youth and adolescence 17:117–133

22. Rasmussen AR, Wohlfahrt-Veje C, Tefre de Renzy-Martin K, Hagen CP, Tinggaard J, Mouritsen A, et al. (2015) Validity of self-assessment of pubertal maturation. Pediatrics 135:86–93

23. Hodes RJ, Insel TR, Landis SC, Research NBfN (2013) The NIH toolbox: setting a standard for biomedical research. Neurology 80:S1–S1

24. Gershon RC, Wagster MV, Hendrie HC, Fox NA, Cook KF, Nowinski CJ (2013) NIH toolbox for assessment of neurological and behavioral function. Neurology 80:S2–S6

25. Acker W (1982) A computerized approach to psychological screening—the Bexley-Maudsley Automated Psychological Screening and the Bexley-Maudsley Category sorting test. International Journal of Man-Machine Studies 17:361–369

26. Moore A, Lewis B, Nixon SJ (2025) Mental rotational skills from pre to mid-adolescence: What a novel test tells us about skill development. Neuropsychology

27. Luciana M, Bjork JM, Nagel BJ, Barch DM, Gonzalez R, Nixon SJ, et al. (2018) Adolescent neurocognitive development and impacts of substance use: Overview of the adolescent brain cognitive development (ABCD) baseline neurocognition battery. Developmental cognitive neuroscience 32:67–79

28. Kentistou KA, Kaisinger LR, Stankovic S, Vaudel M, Mendes de Oliveira E, Messina A, et al. (2024) Understanding the genetic complexity of puberty timing across the allele frequency spectrum. Nature genetics 56:1397–1411

29. Hollis B, Day FR, Busch AS, Thompson DJ, Soares ALG, Timmers PR, et al. (2020) Genomic analysis of male puberty timing highlights shared genetic basis with hair colour and lifespan. Nature communications 11:1536

30. Hatoum AS, Morrison CL, Mitchell EC, Lam M, Benca-Bachman CE, Reineberg AE, et al. (2023) Genome-wide association study shows that executive functioning is influenced by GABAergic processes and is a neurocognitive genetic correlate of psychiatric disorders. Biological psychiatry 93:59–70

31. Burgess S, Thompson SG (2015) Multivariable Mendelian randomization: the use of pleiotropic genetic variants to estimate causal effects. American journal of epidemiology 181:251–260

32. Burgess S, Davies NM, Thompson SG (2016) Bias due to participant overlap in two[sample Mendelian randomization. Genetic epidemiology 40:597–608

33. Hu X, Zhao J, Lin Z, Wang Y, Peng H, Zhao H, et al. (2022) Mendelian randomization for causal inference accounting for pleiotropy and sample structure using genome-wide summary statistics. Proceedings of the National Academy of Sciences 119:e2106858119

34. Baxendale S (2024) The impact of suppressing puberty on neuropsychological function: a review. Acta Paediatrica 113:1156–1167

35. Friedman NP, Robbins TW (2022) The role of prefrontal cortex in cognitive control and executive function. Neuropsychopharmacology 47:72–89

36. Fuhrmann D, Knoll LJ, Blakemore S-J (2015) Adolescence as a sensitive period of brain development. Trends in cognitive sciences 19:558–566

37. Piekarski DJ, Johnson CM, Boivin JR, Thomas AW, Lin WC, Delevich K, et al. (2017) Does puberty mark a transition in sensitive periods for plasticity in the associative neocortex? Brain research 1654:123–144

38. Vijayakumar N, Youssef GJ, Allen NB, Anderson V, Efron D, Hazell P, et al. (2021) A longitudinal analysis of puberty[related cortical development. NeuroImage 228:117684

39. Vijayakumar N, Whittle S, Silk TJ (2023) Corticolimbic connectivity mediates the relationship between pubertal timing and mental health problems. Psychological Medicine 53:7655–7665

40. Auchus RJ, Rainey WE (2004) Adrenarche–physiology, biochemistry and human disease. Clinical endocrinology 60:288–296

