## Supplemental Material for "Puberty Timing and Cognitive Functioning: Insights from the Adolescent Brain Cognitive Development (ABCD) Study and Mendelian Randomization"

### Overview

### Supplementary Methods 1 – ABCD sample analyses

Study Sample

Supplementary Figure S1 - Participant Selection and Final Sample Composition.

Exposure: Puberty Timing

Supplementary Figure S2 – Puberty Timing, PDS Scores and Age in Females at Baseline.

Supplementary Figure S3 – Puberty Timing, PDS Scores and Age in Males at Baseline.

Outcome: Cognitive Measures

Supplementary Figure S4 – NIH Toolbox® Scores by Timepoint.

Supplementary Figure S5 – RAVLT Scores by Timepoint.

Supplementary Figure S6 – LMT Scores by Timepoint.

Covariates and Missingness

Supplementary Figure S7 – Directed Acyclic Graph to Identify a Minimal Adjustment Set

Supplementary Table S1 – Comparison of Included and Excluded Subjects.

Supplementary Table S2 – Comparison of Subjects With and Without Available Birthweight Data.

Supplementary Table S3 – Comparison of Subjects With and Without Available Family Income Class.

Supplementary Table S4 – Comparison of the distribution of observed and imputed family income.

Supplementary Figure S8 – Comparison of the distribution of observed and imputed birthweights.

Linear Mixed Models - Assumptions

Supplementary Table S5 – Variance Inflation Factors per Model to assess Multicollinearity.

Supplementary Figure S9 – QQ Plots to assess normal distribution of residuals in females.

Supplementary Figure S10 – QQ Plots to assess normal distribution of residuals in males.

Supplementary Table S6 – GAMM Results evaluating a smoothed term for ‘Puberty Timing’.

Supplementary Figure S11 – Linear Effect of Puberty Timing on Cognitive Tasks in Females.

Supplementary Figure S12 – Linear Effect of Puberty Timing on Cognitive Tasks in Males.

Sensitivity Analysis

Supplementary Figure S13 – Puberty Timing, PDS Scores and Age in Males at 2-Year Follow-Up.

Supplementary Table S7. Sensitivity Analysis: Effect of Male Puberty Timing at 2-Year Follow-Up on cognitive outcomes.

### Supplementary Methods 2 – Mendelian Randomization analyses

### Supplementary Results 2 – Results of Mendelian Randomization analyses

Age at Menarche

Supplementary Figure S14 – Scatter Plot – Age at Menarche.

Supplementary Figure S15 – Funnel Plot – Age at Menarche.

### Supplementary Figure S16 – MR-APSS Results.

### Supplementary Figure S17 – Reverse Mendelian Randomization analysis.

Male Puberty Timing

Supplementary Figure S18 – Scatter Plot – Male Puberty Timing.

### Supplementary Figure S19 – Funnel Plot – Male Puberty Timing.

**Supplementary References**

### Supplementary Methods 1 - ABCD sample analyses

*Study Sample – Inclusion and Exclusion Criteria*

Subjects were excluded if they lacked basic demographic information (e.g., age or race/ethnicity), had missing baseline pubertal development information, exhibited incongruent sex information (puberty questionnaire versus sex assigned at birth), had implausible Body Mass Index (BMI) values, had congenital sex variations, or were missing baseline cognitive data (Supplementary Figure 1). These criteria resulted in a final sample of N = 10,174 (85.7%) subjects (Table 1). Excluded subjects were more likely to be from ethnic minorities or low-income families (Supplementary Table S1).

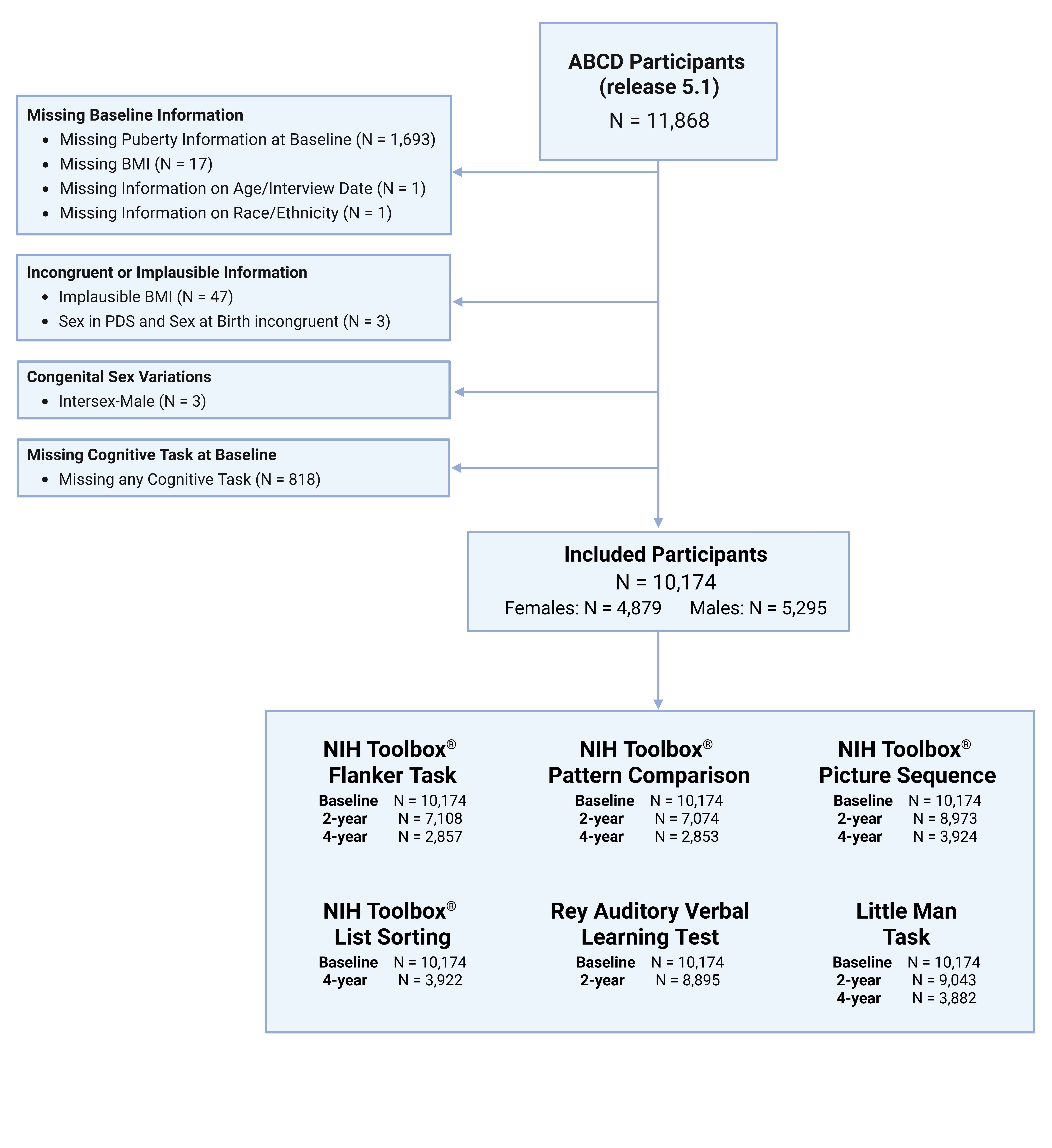
**Supplementary Figure S1 - Participant Selection and Final Sample Composition.** This figure illustrates the inclusion/exclusion criteria used to derive the final study sample.

*Exposure: Puberty Timing*

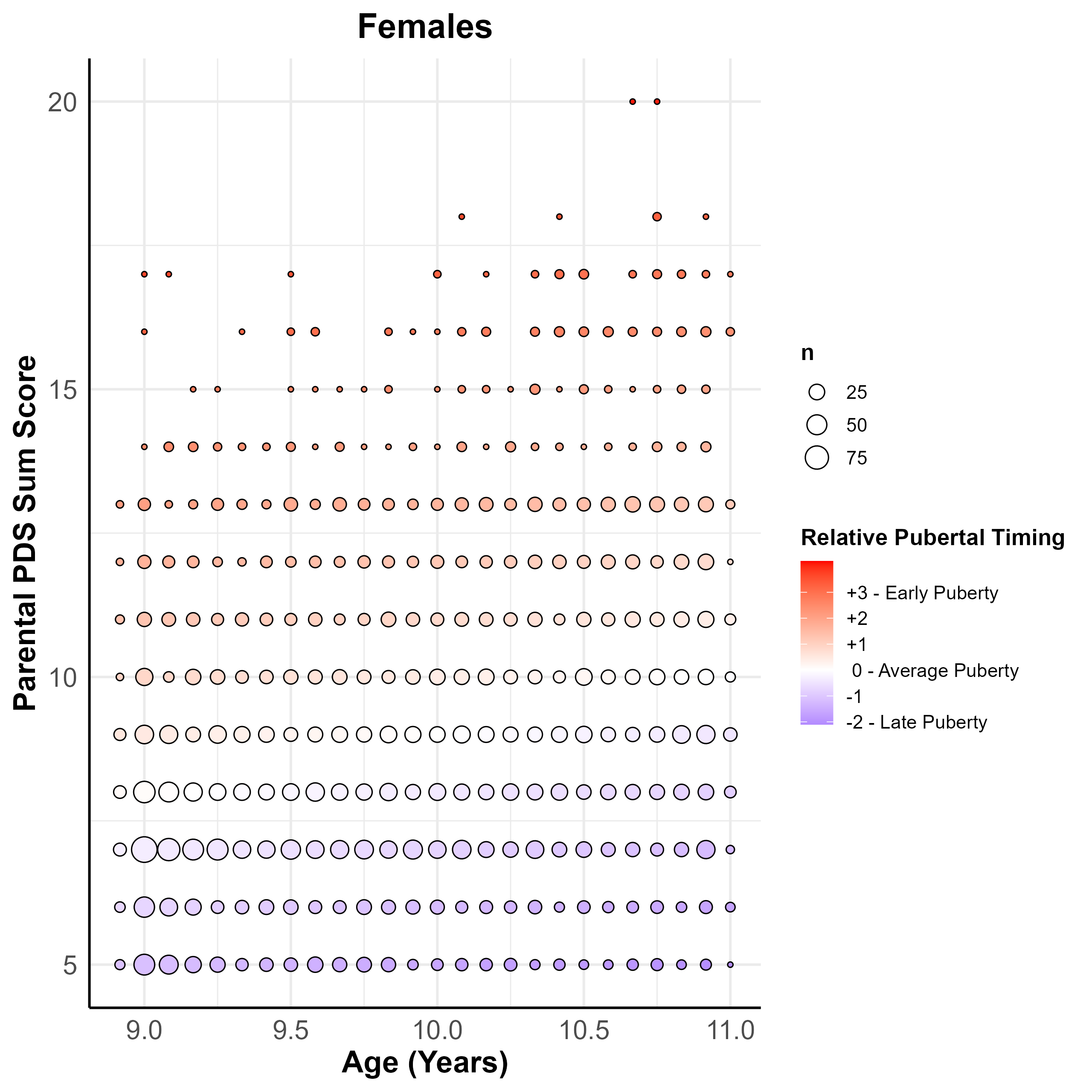

**Supplementary Figure S2 – Puberty Timing, PDS Scores and Age in Females at Baseline.** Puberty timing was operationalized by regressing the sum scores of the Pubertal Development Scale (PDS, [1] on age within each sex and saving the standardized residuals as a measure of Puberty Timing. This plot depicts the resulting relationship between Puberty Timing (red – positive standardized residuals, meaning earlier than average puberty timing), Age (Y-axis, in years) and PDS sum scores (possible ranges: 5 to 20). To account for the large number of overlapping subjects per values, points were sized by the number of participants at each (Age, PDS Score) combination.

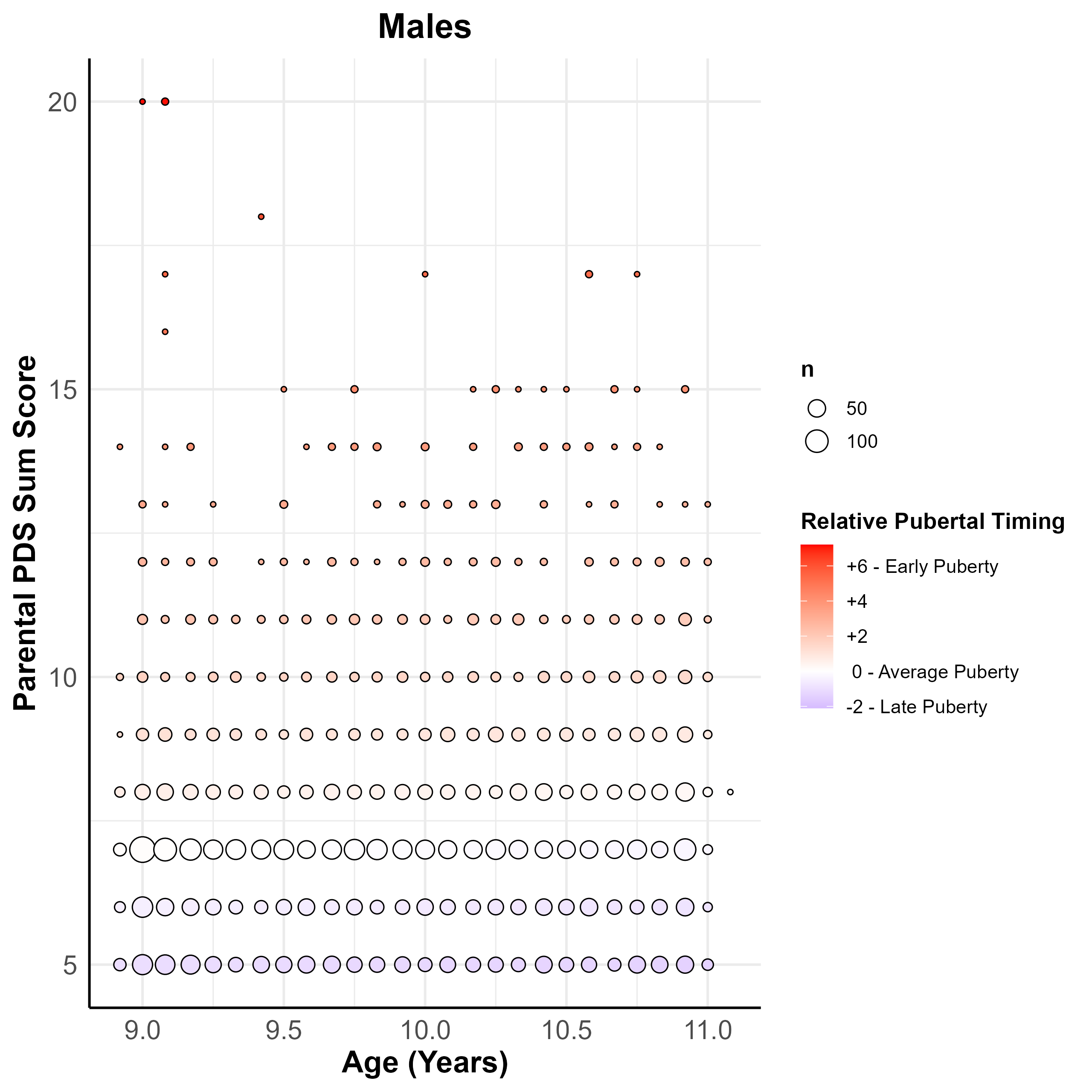

**Supplementary Figure S3 – Puberty Timing, PDS Scores and Age in Males at Baseline.** Puberty timing was operationalized by regressing the sum scores of the Pubertal Development Scale (PDS, [1] on age within each sex and saving the standardized residuals as a measure of Puberty Timing. This plot depicts the resulting relationship between Puberty Timing (red – positive standardized residuals, meaning earlier than average puberty timing), Age (Y-axis, in years) and PDS sum scores (possible ranges: 5 to 20). To account for the large number of overlapping subjects per values, points were sized by the number of participants at each (Age, PDS Score) combination.

*Outcome: Cognitive Measures*

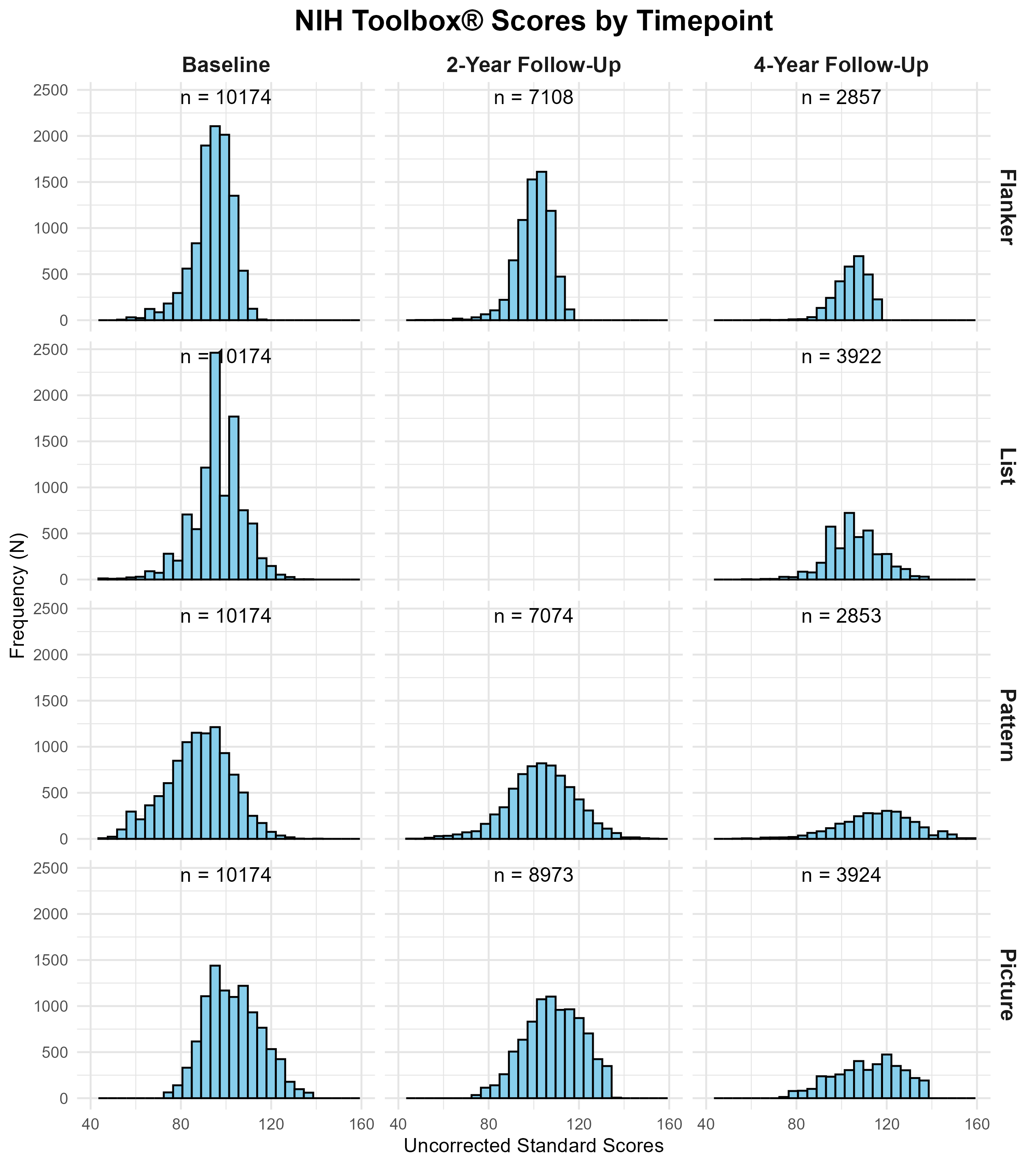
**Supplementary Figure S4 – NIH Toolbox® Scores by Timepoint.** This plot depicts the number of observed uncorrected raw scores (e.g. not corrected for age or sex) per timepoint and measure. Flanker = NIH Toolbox® Flanker Inhibitory Control and Attention Test; List = NIH Toolbox® List Sorting Working Memory Test. Pattern = NIH Toolbox® Pattern Comparison Processing Speed Test; Picture = NIH Toolbox® Picture Sequence Memory Test. Please note that for the NIH Toolbox® List Sorting Task, no observations were available at the 2-Years Follow Up, thus these data is not presented. Uncorrected standard raw scores represent task performance with regard to a normative reference sample with a mean of 100 and a standard deviation of 15 without further adjustments.

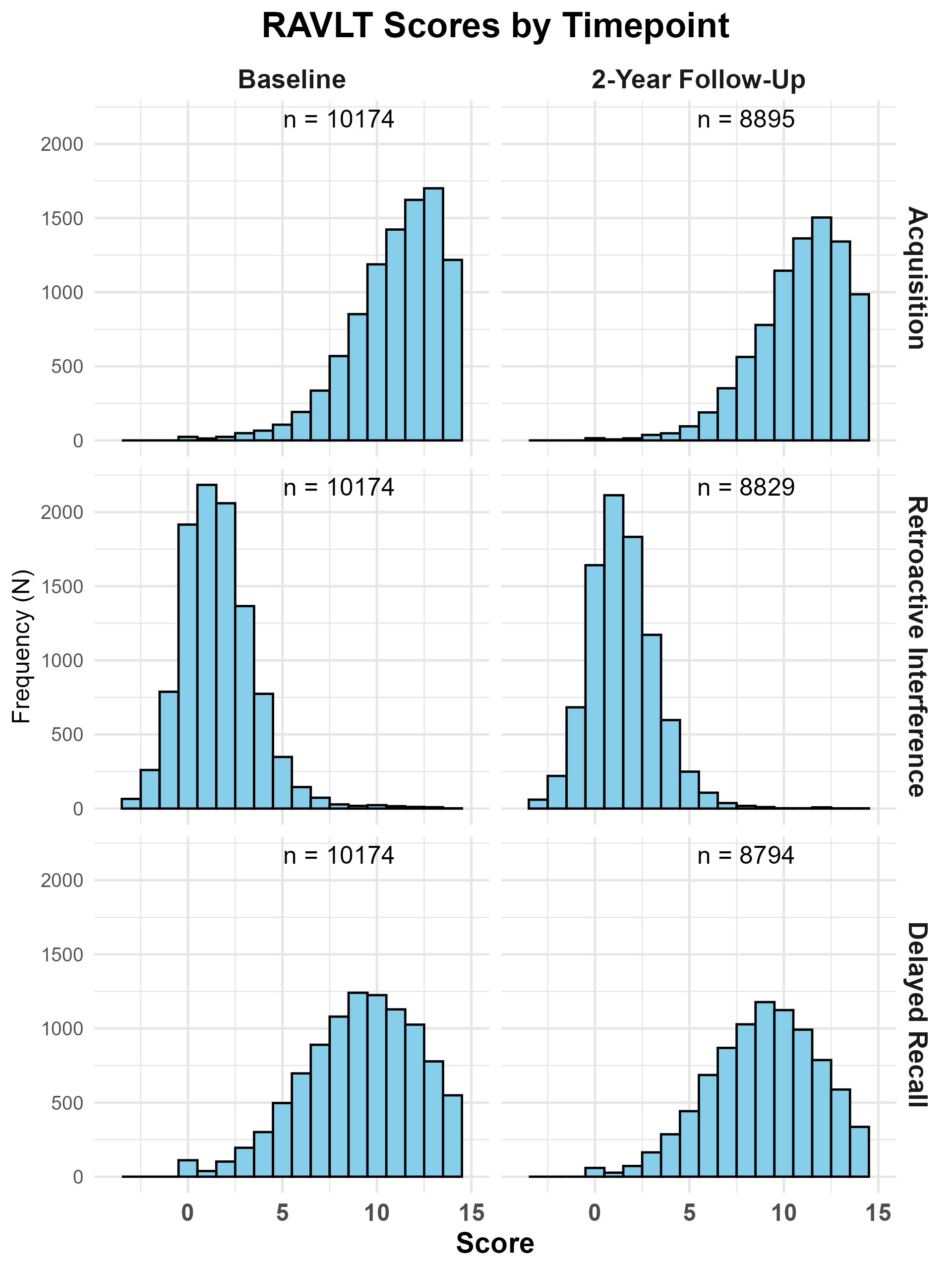

**Supplementary Figure S5 – RAVLT Scores by Timepoint.** Performance of the Rey Auditory Verbal Learning Test (RAVLT) per timepoint. The given score represents the number of recalled items of List A after the last learning trial (RAVLT – Acquisition), the number of learned items (= Acquisition) minus the number of recalled items after presentation of the distractor list (RAVLT - Retroactive Interference), and the number of items recalled after a delay of ~ 30 minutes (RAVLT – Delayed Recall).

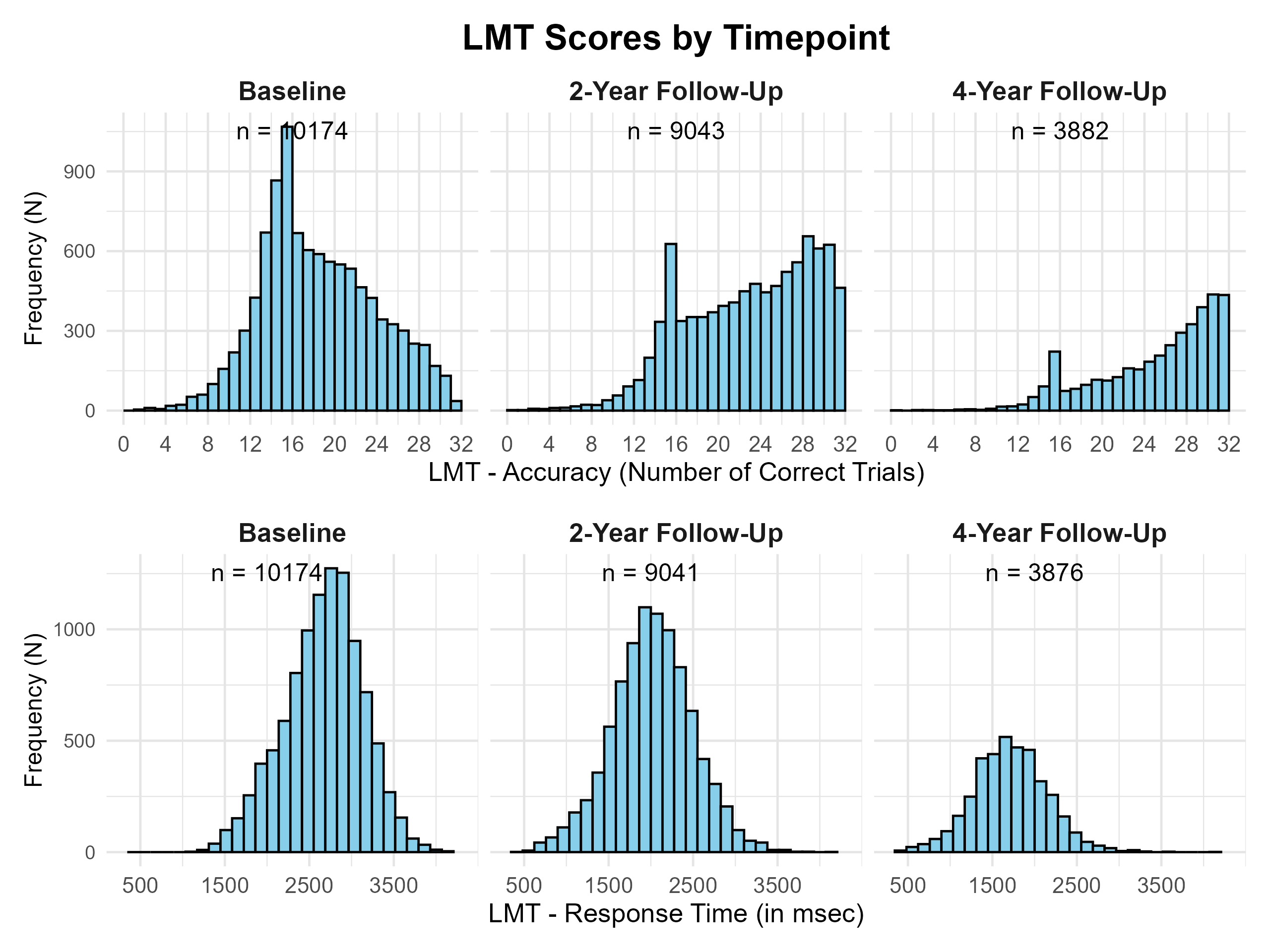
**Supplementary Figure S6 – LMT Scores by Timepoint.** This plot depicts the accuracy (number of correct answers) and response time (response time in msec = milliseconds for correct answers) of the little man task (LMT) per timepoint.

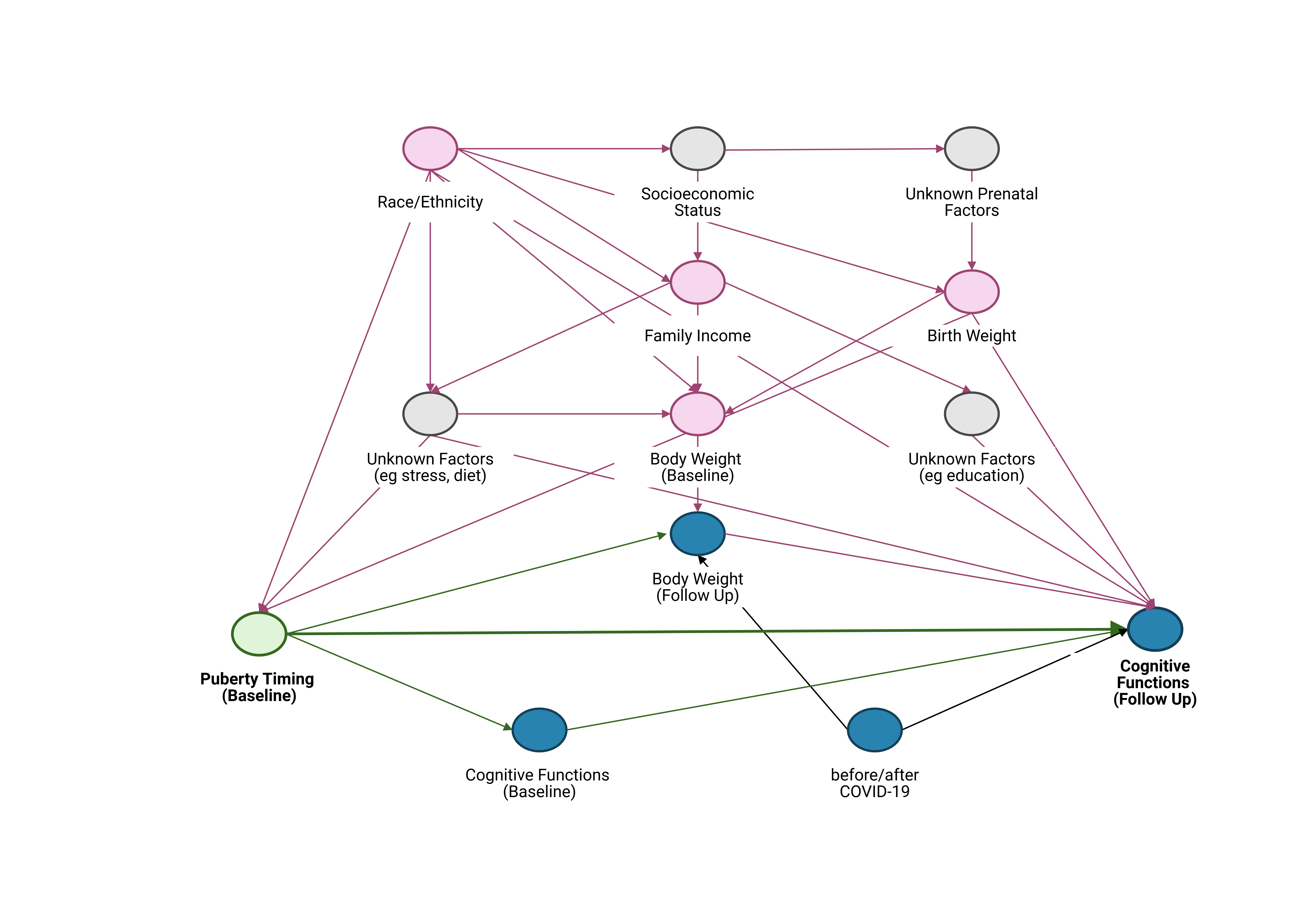

**Supplementary Figure S7 – Directed Acyclic Graph to Identify a Minimal Adjustment Set.** To examine the relationship between potential confounders, a directed acyclic graph (DAG) was constructed using the online tool DAGitty [2]. The DAG was used to identify the minimally sufficient set of adjustment variables for estimating the total causal effect of the exposure (puberty timing at baseline) on the outcome (performance in cognitive tests) [3]. In the graph, the exposure variable is shown in green (highlighted in bold). The outcome variable (highlighted in bold), along with its ancestors, is shown in blue. Observed confounding variables are displayed in red, and unobserved variables in gray. Although multiple cognitive outcome measures were analyzed, a single DAG was applied under the assumption that the causal structure of the relationships is consistent across cognitive domains. We assumed that family income influences both puberty timing—potentially through mechanisms such as stress, nutrition, or body weight [4, 5]—and cognitive performance, for example, through familial stress or the family's ability to provide academic support such as private tutoring [6, 7]. Regarding birthweight, we based our assumptions on findings linking lower birthweight to both reduced executive functioning—particularly working memory [8]—and earlier puberty timing [9]. For body weight, the assumptions relied on extensive evidence connecting higher body weight to earlier pubertal maturation [5] and a bidirectional relationship with executive functioning [10]. For the purposes of our model, we considered the pathway from body weight to cognitive functioning as the more relevant confounding direction. With regard to cognitive functioning at baseline (i.e., the time point at which puberty timing was measured), we assumed that puberty timing may already exert an influence on cognitive performance. Consequently, baseline cognitive functioning could act as a mediator in the pathway from puberty timing at baseline to cognitive performance at a later follow-up. As our aim was to estimate the total causal effect of puberty timing on subsequent cognitive performance, we did not adjust for baseline cognitive functioning, in order to avoid conditioning on a potential mediator. Based on these assumptions, the minimal sufficient adjustment sets for estimating the total effect of Puberty Timing (Baseline) on Cognitive Functions (Follow-Up) is: Birth Weight, Body Weight (Baseline), Family Income, Race/Ethnicity.

*Covariates*

To control for body weight, the BMI standard deviation score was calculated based on weight and height. For the measurement of weight, participants were instructed to remove extra clothing (e.g., jackets), shoes, and empty their pockets before measurement. Each participant was weighed twice; if the two readings differed by more than 0.1 lb, a third measurement was taken. The final weight (in pounds) was determined by averaging the two closest measurements. If the third measurement was equidistant from the first two, all three were averaged. Height: Standing height (in inches) was measured twice; if discrepancies occurred, a third measure was obtained. The two closest readings were averaged, or, if the third was equidistant, all three were averaged. Weight and height were then converted to kilograms and centimeters using the measurements package (v1.5.1). Standard deviation scores (BMI-SDS) were obtained using the CDC reference growth charts [11] with the ‘childsds’ package (version 0.9.8) based on age and sex. In 13 subjects, information on height and/or weight was missing. For 53 participants, weight and/or height information seemed implausible, leading to BMI values from 2.1 to 3017.3kg/m^2^, leading to extreme or non-detectable BMI-SDS values. Therefore, participants with BMI-SDS values below -4 or above +8 or missing BMI values were excluded (N = 62, 0.5%).

*Missingness*

We decided to exclude patients with missing values in the baseline exposure (PDS score) or outcome variables (cognitive measures) of interest, and subjects with missing or implausible information for covariates (see Figure 1), resulting in a total of 10,175 included subjects out of the total sample of 11,868 subjects (85.7%). Excluded subjects had similar age than included subjects, but were more likely to belong to an ethnic minority and had lower family income (see Supplementary Table S1 below). Given the relatively high percentage (> 1%) of subjects with missing information on birthweight and family income, we decided to impute missing values for these two variables to avoid exclusion of a relevant number of subjects due to missing covariate information. leading to BMI values from 2.1 to 3017.3kg/m2, leading to extreme or non-detectable BMI-SDS values.

| **Supplementary Table S1. Comparison of Included and Excluded Subjects.** | | | | |
| --- | --- | --- | --- | --- |
| **Characteristics** | | **Included**  **(N = 10,174)** | **Excluded**  **(N = 1,694)** | **p values ^a^**  **t-test/Χ²-test** |
| **Age (Years), mean (SD)** | | 9.91 (0.63) | 9.92 (0.62) | 0.98 |
| **Sex, N (%)** | |  |  |  |
|  | **Female** | 5,295 (52.0) | 893 (52.7) | < 0.001 |
|  | **Male** | 4,879 (48.0) | 798 (47.1) |  |
|  | **Intersex-Male** | 0 (0.0) | 3 (0.2) |  |
| **Race/Ethnicity, N (%)** | |  |  |  |
|  | White | 5,520 (54.3) | 653 (38.6) | < 0.001 |
|  | Black | 1,371 (13.5) | 413 (24.4) |  |
|  | Hispanic | 1,992 (19.6) | 418 (24.7) |  |
|  | Asian | 224 (2.2) | 28 (1.7) |  |
|  | Other/Multiethnic | 1,067 (10.5) | 181 (10.7) |  |
| **Family Income (past year) ^a^, N (%)** | |  |  |  |
|  | < $5000 | 342 (3.4) | 147 (8.7) | < 0.001 |
|  | $5,000 - $11,999 | 373 (3.7) | 112 (6.6) |  |
|  | $12,000 - $15,999 | 244 (2.4) | 83 (4.9) |  |
|  | $16,000 - $24,999 | 495 (4.9) | 115 (6.8) |  |
|  | $25,000 - $34,999 | 602 (5.9) | 142 (8.4) |  |
|  | $35,000 - $49,999 | 855 (8.4) | 178 (10.5) |  |
|  | $50,000 - $74,999 | 1,410 (13.9) | 225 (13.3) |  |
|  | $75,000 - $99,999 | 1,468 (14.4) | 199 (11.7) |  |
|  | $100,000 - $199,999 | 3,181 (31.3) | 367 (21.7) |  |
|  | > $200,000 | 1,204 (11.8) | 126 (7.4) |  |

Abbreviations: N = Number, SD = Standard deviation.

a. For categorical data (sex, race/ethnicity, family income class), Chi-squared tests were calculated. For numeric data (age), a t-test was calculated.

Birth Weight – For birth weight, parents reported their child’s weight in pounds and additional ounces, which was then converted to kilograms. If the weight in pounds was unknown (coded as 999) or missing, the birth weight was treated as missing. If the ounces data were missing, only the pounds value was converted and used. In 517 cases (4.4%), birth weight was missing and subsequently imputed (see rationale and methods below).

| **Supplementary Table S2. Comparison of Subjects With and Without Available Birthweight Data** | | | | |
| --- | --- | --- | --- | --- |
| **Characteristics** | | **Available**  **(N = 11,351)** | **Missing**  **(N = 517)** | **p values ^a^**  **t-test/Χ²-test** |
| **Age (Years), mean (SD)** | | 9.91 (0.63) | 9.93 (0.60) | 0.482 |
| **Sex, N (%)** | |  |  |  |
|  | **Female** | 5,915 (52.1) | 273 (52.8) | 0.892 |
|  | **Male** | 5,433 (47.9) | 244 (47.2) |  |
|  | **Intersex-Male** | 3 (0.0) | 0 (0.0) |  |
| **Race/Ethnicity, N (%)** | |  |  |  |
|  | White | 6,024 (53.1) | 149 (28.8) | < 0.001 |
|  | Black | 1,637 (14.4) | 147 (28.4) |  |
|  | Hispanic | 2,287 (20.1) | 123 (23.8) |  |
|  | Asian | 220 (1.9) | 32 (6.2) |  |
|  | Other/Multiethnic | 1,182 (10.4) | 66 (12.8) |  |
| **Family Income (past year) ^a^, N (%)** | |  |  |  |
|  | < $5000 | 454 (4.1) | 35 (6.8) | < 0.001 |
|  | $5,000 - $11,999 | 467 (4.1) | 18 (3.5) |  |
|  | $12,000 - $15,999 | 304 (2.7) | 23 (4.5) |  |
|  | $16,000 - $24,999 | 567 (5.0) | 43 (8.3) |  |
|  | $25,000 - $34,999 | 698 (6.1) | 46 (8.9) |  |
|  | $35,000 - $49,999 | 973 (8.6) | 60 (11.6) |  |
|  | $50,000 - $74,999 | 1,565 (13.8) | 70 (13.5) |  |
|  | $75,000 - $99,999 | 1,599 (14.1) | 68 (13.2) |  |
|  | $100,000 - $199,999 | 3,441 (30.3) | 107 (20.7) |  |
|  | > $200,000 | 1,283 (11.3) | 47 (9.1) |  |

Abbreviations: N = Number, SD = Standard deviation.

a. For categorical data (sex, race/ethnicity, family income class), Chi-squared tests were calculated. For numeric data (age), a t-test was calculated.

Family Income – For family income, parents were given a set of ten income classes (see Supplementary Table S1 for boundaries used) asked at the baseline visit: “*Which of these categories best describes your TOTAL COMBINED FAMILY INCOME for the past 12 months? This should include income (before taxes and deductions) from all sources, wages, rent from properties, social security, disability and/or veteran's benefits, unemployment benefits, workman's compensation, help from relative (include child payments and alimony), and so on*”. For two subjects, information was missing. For N = 511 subjects, parents refused to answer the question (coded as 777). For N = 504, parents stated that they did not know the information (coded as 999), leading to a total of N = 1,017 (8.6%) missing values.

| **Supplementary Table S3. Comparison of Subjects With and Without Available Family Income Class** | | | | |
| --- | --- | --- | --- | --- |
| **Characteristics** | | **Available**  **(N = 10,851)** | **Missing**  **(N = 1,017)** | **p values ^a^**  **t-test/Χ²-test** |
| **Age (Years), mean (SD)** | | 9.91 (0.62) | 9.93 (0.62) | 0.548 |
| **Sex, N (%)** | |  |  |  |
|  | **Female** | 5,642 (52.0) | 546 (53.7) | 0.176 |
|  | **Male** | 5,207 (48.0) | 470 (46.2) |  |
|  | **Intersex-Male** | 2 (0.0) | 1 (0.0) |  |
| **Race/Ethnicity, N (%)** | |  |  |  |
|  | White | 5,878 (54.2) | 295 (29.0) | < 0.001 |
|  | Black | 1,511 (13.9) | 273 (26.8) |  |
|  | Hispanic | 2,103 (19.4) | 307 (30.2) |  |
|  | Asian | 217 (2.0) | 35 (3.4) |  |
|  | Other/Multiethnic | 1,141 (10.5) | 107 (10.5) |  |

Abbreviations: N = Number, SD = Standard deviation.

a. For categorical data (sex, race/ethnicity), Chi-squared tests were calculated. For numeric data (age), a t-test was calculated.

*Imputation of missing family income classes and birthweight with MICE*

Missing data can be categorized as Missing Completely at Random (MCAR), which implies that missingness is entirely random and unrelated to observed or unobserved variables; Missing at Random (MAR), where missingness is related only to observed variables; or Missing Not at Random (MNAR), where missingness is related to unobserved data. As shown in Supplementary Tables S2 and S3, missingness for birthweight and family income was associated with basic demographic variables, making MCAR unlikely. Although we could not directly distinguish MAR from MNAR (because unobserved data were unattainable), we employed Multiple Imputation by Chained Equations (MICE; mice package version 3.17.0, [12]) under the MAR assumption. For family income (categorical), we used polytomous regression with five iterations, including site-ID and race/ethnicity as predictors. Supplementary Table S4 compares the distribution of observed and imputed family income classes. For birthweight (numerical), we used predictive mean matching with five iterations, including site-ID, sex, prematurity, family income, and race/ethnicity as predictors. Supplementary Figure S8 compares the distribution of observed and imputed birthweight.

**Supplementary Table S4. Comparison of the distribution of observed and imputed family income.**

| **Characteristics** | | **Observed**  **(N = 10,851)** | **Imputed**  **(N = 1,017)** |
| --- | --- | --- | --- |
| **Family Income (past year) ^a^, N (%)** | |  |  |
|  | < $5000 | 417 (3.8) | 72 (7.1) |
|  | $5,000 - $11,999 | 421 (3.9) | 64 (6.3) |
|  | $12,000 - $15,999 | 273 (2.5) | 54 (5.3) |
|  | $16,000 - $24,999 | 523 (4.8) | 87 (8.6) |
|  | $25,000 - $34,999 | 654 (6.0) | 90 (8.8) |
|  | $35,000 - $49,999 | 934 (8.6) | 99 (9.7) |
|  | $50,000 - $74,999 | 1,498 (13.8) | 137 (13.5) |
|  | $75,000 - $99,999 | 1,570 (14.5) | 97 (9.5) |
|  | $100,000 - $199,999 | 3,311 (30.5) | 237 (23.3) |
|  | > $200,000 | 1,250 (11.5) | 80 (7.8) |

Abbreviations: N = Number.

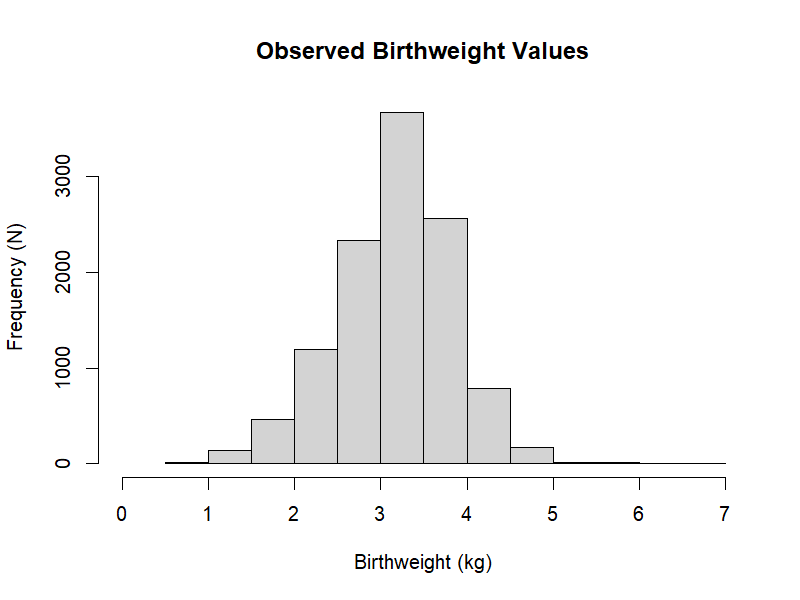

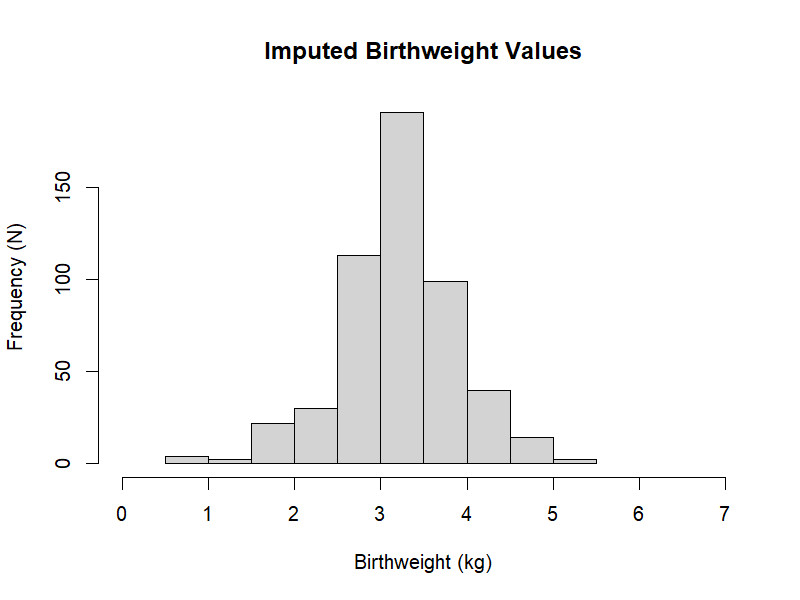

**Supplementary Figure S8 – Comparison of the distribution of observed and imputed birthweights.**

*Linear Mixed Models – Assumptions*

Multicollinearity: To assess multicollinearity among predictors, we computed generalized variance inflation factors (GVIFs). These were derived from linear models that included only fixed effects, excluding random terms. To facilitate comparison across categorical variables with differing numbers of levels, we scaled the GVIFs using the transformation GVIF^1/(2df)^, and interpreted the squared values of these adjusted GVIFs (i.e., GVIF^1/df^). A squared adjusted GVIF exceeding a threshold of 4–5 was considered indicative of potentially problematic multicollinearity warranting further examination (for a discussion of this threshold, see [13]). None of the covariates approached this threshold, suggesting that problematic multicollinearity was unlikely (see Supplementary Table S5).

**Supplementary Table S5. Variance Inflation Factors per Model to assess Multicollinearity.**

|  | | **Females** | | **Males** | |
| --- | --- | --- | --- | --- | --- |
| **Variable** | | GVIF | GVIF^1/df^ | GVIF | GVIF^1/df^ |
|  | Pubertal Timing | 1.35 | 1.35 | 1.17 | 1.17 |
|  | BMI-SDS | 1.27 | 1.28 | 1.12 | 1.12 |
|  | Birthweight | 1.03 | 1.04 | 1.03 | 1.03 |
|  | Race/Ethnicity | 1.51 | 1.11 | 1.47 | 1.10 |
|  | Family Income Class | 1.43 | 1.04 | 1.40 | 1.04 |

Abbreviation: BMI-SDS = Body Mass Index – Standard Deviation Scores; GVIF = Generalized variance inflation factors. GVIF^1/df^ = squared adjusted GVIF.

Normality of residuals: To assess the assumption of normally distributed residuals in the linear mixed-effects models, Quantile–Quantile (QQ) plots were generated for each of the nine cognitive outcome measures across all timepoints, stratified by sex. The plots compared the quantiles of the standardized residuals from the sample to the theoretical quantiles expected under a normal distribution (see Supplementary Figures S9 and S10). Visual inspection revealed only minor deviations from normality for most variables, which were considered negligible given the large sample size of the present study.

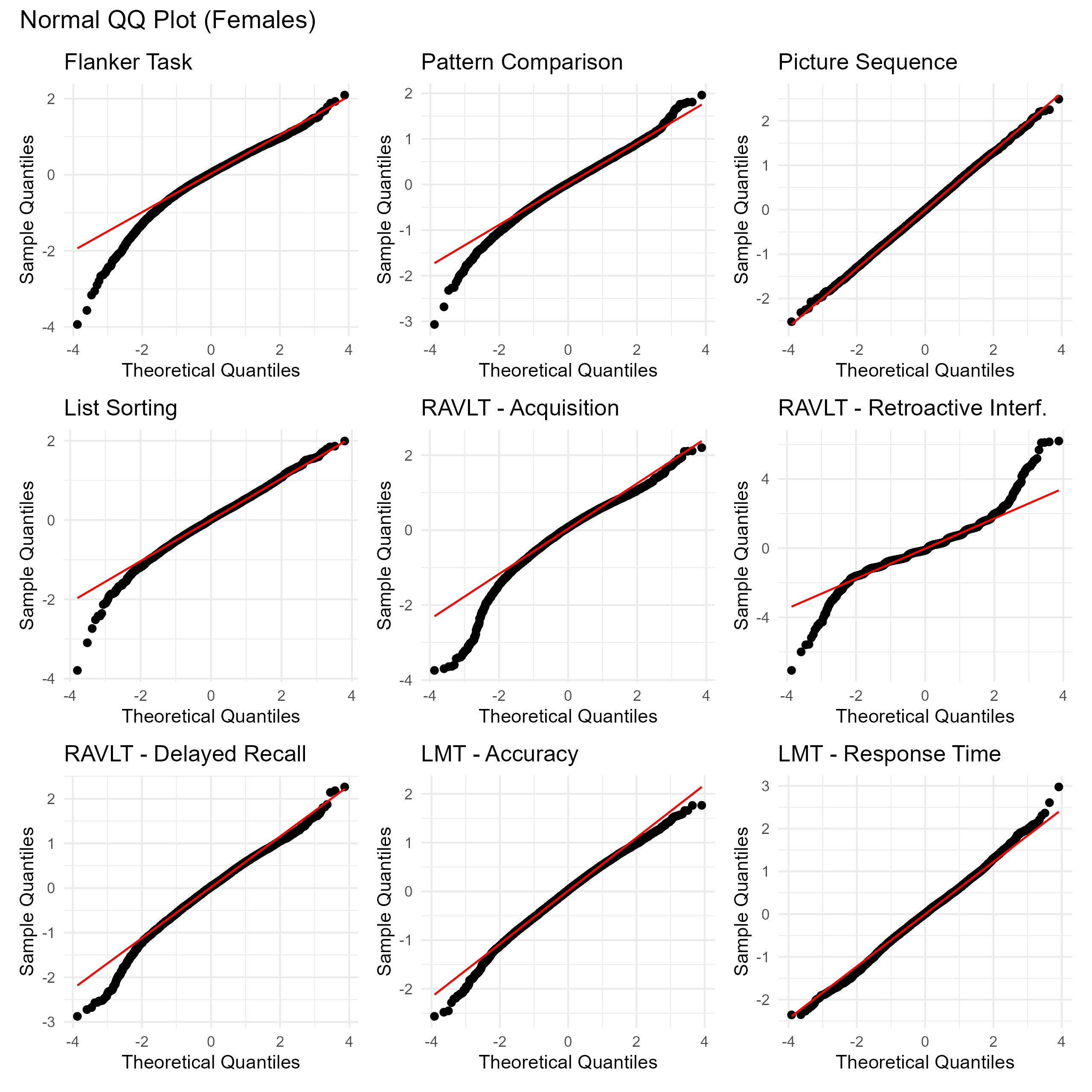
**Supplementary Figure S9 – QQ Plots to assess normal distribution of residuals in females.**

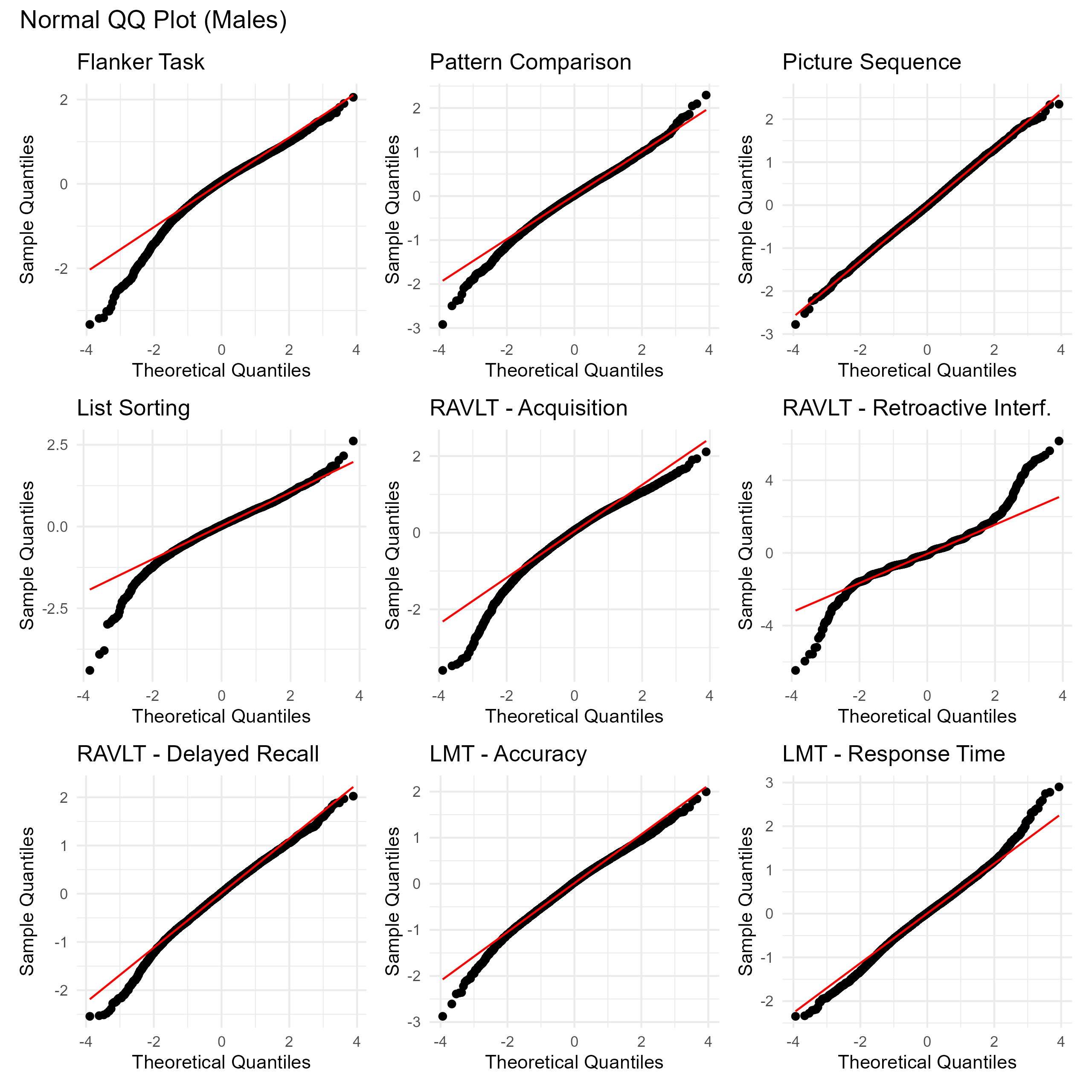
**Supplementary Figure S10 – QQ Plots to assess normal distribution of residuals in males.**

Non-linearity: To evaluate the suitability of linear models, we fitted Generalized Additive Mixed Models (GAMMs) incorporating penalized thin plate regression splines for Puberty Timing across all timepoints (‘Overall’), stratified by sex and adjusted for the same fixed and random effects as in the main analysis. Additionally, standardized cognitive task performance was plotted against binned mean values of Puberty Timing (± standard error of the mean) to visually assess potential non-linear associations in the raw data. The GAMMs with and without a smoothed term were compared with the Akaike Information Criterion (AIC), with a Δ AIC of ≥ 2 was interpreted as a meaningful difference favoring one of these models [14]. In addition, a likelihood ratio test (LRT) was used to compare model performance [15]. Only one model exhibited a Δ AIC ≥ 2, favoring the model with a smoothed term (RAVLT-Acquisition in males, Δ AIC = 3.69), however the LRT comparing the performance of GAMMs with linear vs. the smoothed term for these models remained non-significant (p = 0.134). Visual inspection of the raw data suggested a linear relationship for this task, implying that the observed non-linearity may have been introduced by the inclusion of additional covariates. Given this observation, and in line with our goal of building parsimonious models, we focused our interpretation on the linear effects of Puberty Timing.

**Supplementary Table S6. GAMM Results evaluating a smoothed term for ‘Puberty Timing’.**

|  | | **Females** | | | | **Males** | | | |
| --- | --- | --- | --- | --- | --- | --- | --- | --- | --- |
| **Variable** | | EDF ^a^ | p_smooth_ ^b^ | Δ AIC ^c^ | p_LRT_ ^d^ | EDF ^a^ | p_smooth_ ^b^ | Δ AIC ^c^ | p_LRT_ ^d^ |
|  | NIH Toolbox®  Flanker Task | 1.00 | 0.071 | -2.03 | 1.000 | 1.00 | 0.058 | -2.06 | 1.000 |
|  | NIH Toolbox®  Pattern Comparison | 3.09 | 0.375 | -1.53 | 1.000 | 1.58 | 0.755 | -1.90 | 1.000 |
|  | NIH Toolbox®  Picture Sequence | 1.13 | 0.004 | -2.01 | 1.000 | 1.17 | 0.019 | -2.01 | 1.000 |
|  | NIH Toolbox®  List Sorting | 1.14 | 0.010 | -2.02 | 1.000 | 3.27 | < 0.001 | 1.34 | 0.949 |
|  | RAVLT  Acquisition | 1.00 | < 0.001 | -2.00 | 1.000 | **3.22** | **< 0.001** | **3.69** | **0.131** |
|  | RAVLT  Delayed Recall | 1.00 | 0.017 | -2.01 | 1.000 | 2.96 | 0.003 | 1.41 | 0.583 |
|  | RAVLT  Retroactive Interference | 1.00 | 0.819 | -2.00 | 1.000 | 1.45 | 0.583 | -1.87 | 1.000 |
|  | LMT  Accuracy | 2.33 | 0.012 | 0.39 | 0.619 | 1.77 | 0.089 | -1.63 | 1.000 |
|  | LMT  Response Time | 1.00 | 0.004 | -2.02 | 1.000 | 1.00 | 0.959 | -2.03 | 1.000 |

Abbreviation: EDF = Effective Degrees of Freedom; GAMM = Generalized Additive Mixed Models; AIC = Akaike Information Criterion; LRT = Likelihood Ratio Test.

a. An EDF value of 1.00 indicates that the GAMM (Generalized Additive Mixed Model) estimated a linear relationship for "Puberty Timing." Values of EDF greater than 1.00 indicate a non-linear (smoothed) term and were subsequently highlighted.
b. P-values representing the effect of Puberty Timing on each cognitive task, as estimated by a GAMM with a smoothed term (k = 5) for puberty timing (at baseline). The models also included fixed effects family income, race/ethnicity and BMI-SDS, birthweight. Random intercepts were included for participant, family (nested within site), and timepoint.
c. AIC difference between GAMMs with a smoothed versus a linear term for Puberty Timing. A negative Δ AIC indicates a better model fit with a linear term, a positive Δ AIC a better fit with a smoothed term; differences ≥ 2 were interpreted as relevant [14].
d. P-values of the likelihood ratio test (LRT) analyzing whether the model performance of the more complex model (with a smoothed term) is significantly better (based on the AIC) than the simpler model (with a linear term; both models fitted under Maximum Likelihood Estimation). P-values < 0.05 were considered as a statistically significant.

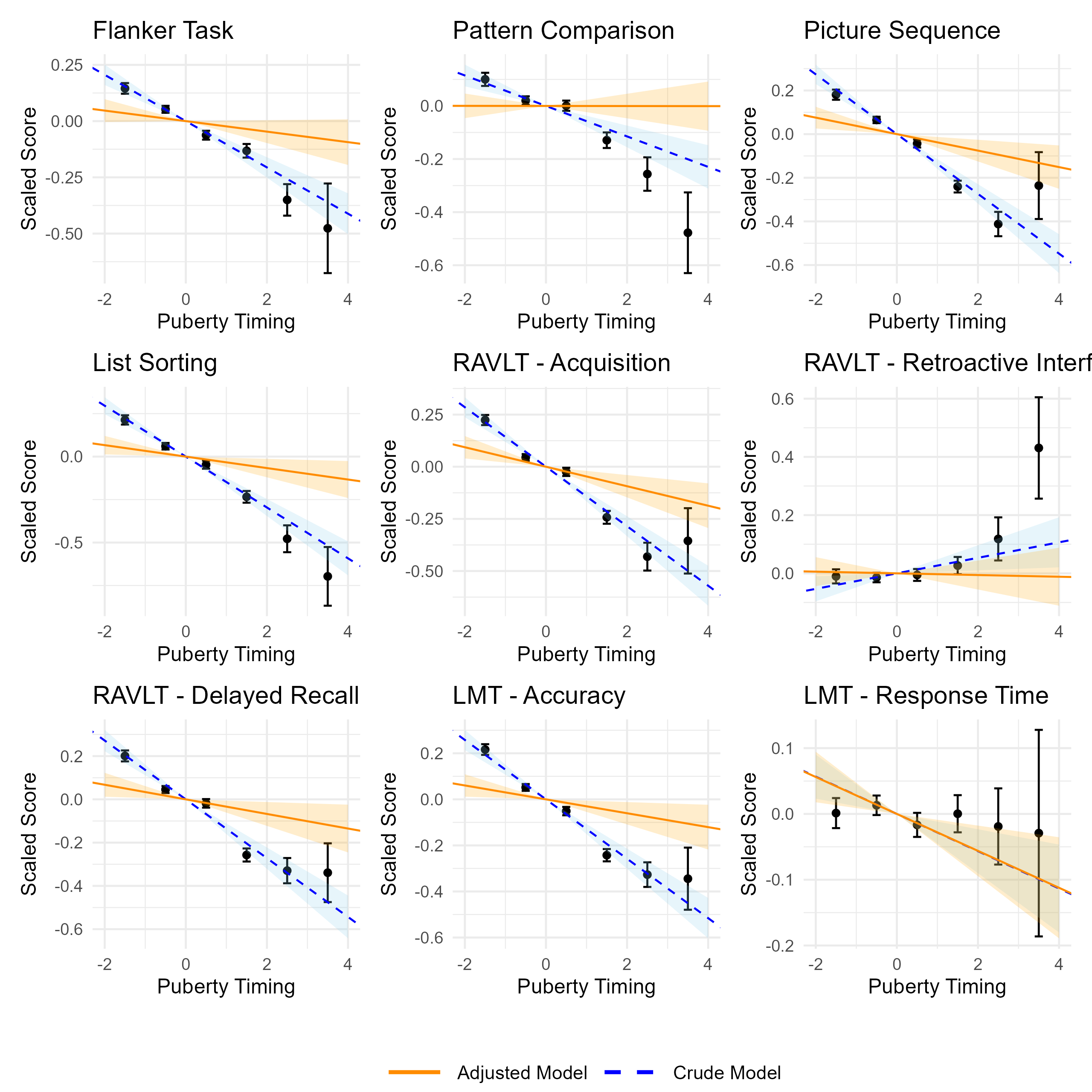
**Supplementary Figure S11 – Linear Effect of Puberty Timing on Cognitive Tasks in Females.** Dots represent the mean standardized performance on each cognitive task (across all timepoints) within each bin of Puberty Timing (e.g., from -2 to -1). Error bars indicate the standard error of the mean. The ‘Crude Model’ (blue, dashed line, with 95% confidence bands) included puberty timing at baseline as a fixed effect and random intercepts for subject, timepoint, and family nested within site (linear mixed models). The ‘Adjusted Model’ (orange, solid line) additionally accounted for BMI-SDS, race/ethnicity, family income, and birthweight.

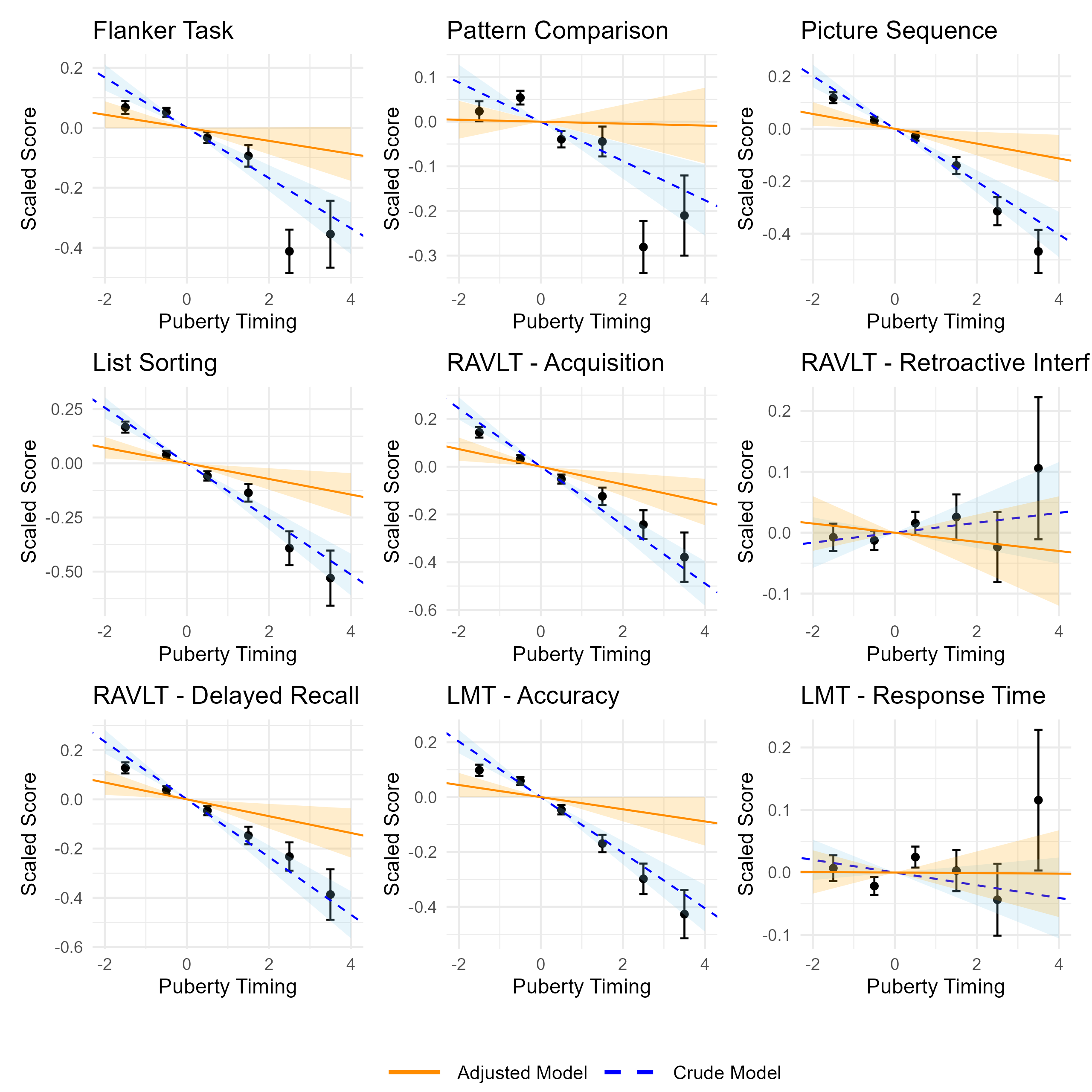

**Supplementary Figure S12 – Linear Effect of Puberty Timing on Cognitive Tasks in Males.** Dots represent the mean standardized performance on each cognitive task (across all timepoints) within each bin of Puberty Timing (e.g., from -2 to -1). Error bars indicate the standard error of the mean. The ‘Crude Model’ (blue, dashed line, with 95% confidence bands) included puberty timing (at baseline) as a fixed effect and random intercepts for subject, timepoint, and family nested within site. The ‘Adjusted Model’ (orange, solid line) additionally accounted for BMI-SDS, race/ethnicity, family income, and birthweight.

Software and packages: All statistical analyses were run on R, version 4.4.2. Linear mixed models were calculated using the *lme4* package, version 1.1.36 [16]. P values for these models were derived using the *lmerTest* package, version 3.1.3 [17]. GAMMs were calculated using *gamm4*, version 0.2.6. GVIF were calculated using the *car* package in R (version 3.1.3; [18])

*Sensitivity Analysis with male puberty timing measured at the 2-year follow-up*

To assess whether the limited variability in male PT at baseline (age 9.9 ± 0.6 years) affected results, we re-ran the analysis using PT measured at the 2-year follow-up (age 12.0 ± 0.7 years).

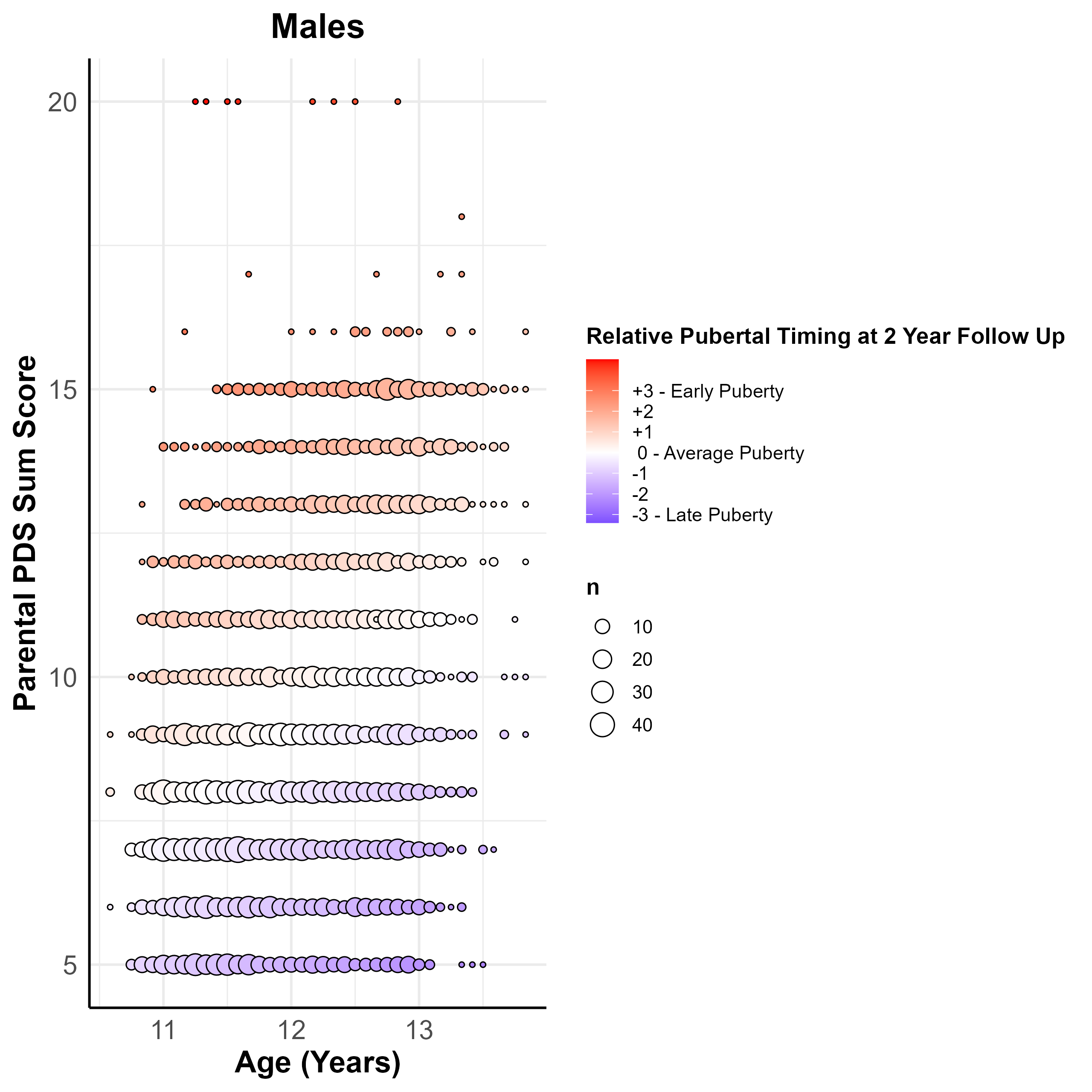

**Supplementary Figure S13 – Puberty Timing, PDS Scores and Age in Males at 2-Year Follow-Up.** Puberty timing was operationalized by regressing the sum scores of the Pubertal Development Scale at the 2-Year Follow-Up (PDS, [1] on age within each sex and saving the standardized residuals as a measure of Puberty Timing. This plot depicts the resulting relationship between Puberty Timing (red – positive standardized residuals, meaning earlier than average puberty timing), Age (Y-axis, in years) and PDS sum scores (possible ranges: 5 to 20). To account for the large number of overlapping subjects per values, points were sized by the number of participants at each (Age, PDS Score) combination.

**Supplementary Table S7. Sensitivity Analysis: Effect of Male Puberty Timing on cognitive outcomes across timepoints.**

| **Cognitive Task (across timepoints)** |  | **Effect of Puberty Timing**  **Measured at Baseline**  (Age 9.9±0.6) | | | | **Effect of Puberty Timing measured at 2-Years**  (Age 12.0±0.7) | | | |
| --- | --- | --- | --- | --- | --- | --- | --- | --- | --- |
|  |  | N_obs_ | β | 95%-CI | p | N_obs_ | β | 95%-CI | p |
| NIH Toolbox®  Flanker Task |  | 10,533 | -0.02 | (-0.04, 0.00) | 0.058 | 10,149 | -0.01 | (-0.03, 0.01) | 0.493 |
| NIH Toolbox®  Pattern Comparison |  | 10,513 | -0.00 | (-0.02, 0.02) | 0.840 | 10,131 | 0.01 | (-0.01, 0.04) | 0.193 |
| NIH Toolbox®  Picture Sequence |  | 12,051 | -0.03 | (-0.05, -0.01) | 0.014 | 11,659 | -0.04 | (-0.07, -0.02) | <0.001 |
| NIH Toolbox®  List Sorting |  | 7,409 | -0.04 | (-0.06, -0.01) | 0.004 | 7,021 | -0.03 | (-0.05, -0.00) | 0.039 |
| RAVLT  Acquisition |  | 9,922 | -0.04 | (-0.06, -0.01) | 0.003 | 9,563 | -0.03 | (-0.05, -0.00) | 0.025 |
| RAVLT  Delayed Recall |  | 9,864 | -0.03 | (-0.06, -0.01) | 0.007 | 9,508 | -0.02 | (-0.04, 0.01) | 0.119 |
| RAVLT  Retroactive Interference |  | 9,880 | -0.01 | (-0.03, 0.01) | 0.511 | 9,522 | -0.02 | (-0.04, 0.01) | 0.159 |
| LMT  Accuracy |  | 12,060 | -0.02 | (-0.04, -0.00) | 0.048 | 11,659 | -0.02 | (-0.04, 0.01) | 0.178 |
| LMT  Response Time |  | 12,056 | -0.00 | (-0.02, 0.02) | 0.959 | 11,654 | -0.02 | (-0.03, 0.00) | 0.069 |

Due to the later onset of puberty in males, this sensitivity analysis examined whether puberty timing at the 2-year follow-up reproduced the main findings. The table shows associations between puberty timing at 2 years and cognitive task performance, estimated using mixed linear models across all timepoints (baseline, 2-year, and 4-year follow-ups). Effect sizes are standardized (SD change in cognition per SD increase in puberty timing). Models were adjusted for BMI-SDS, race/ethnicity, family income, and birthweight, with random intercepts for subject, timepoint, and family nested within site. Abbreviation: N_obs_ = Number of observations.

**Supplementary Methods 2 – Mendelian Randomization analyses**

Information on lead SNPs for “Age at Menarche” and “Male Puberty Timing”—including beta, standard error, effect allele, and effect-allele frequency—was obtained from Kentistou, et al. [19] (Supplemental Table “ST2 | 1080 signals”) for Age at Menarche and from Hollis, et al. [20] (Supplemental Data 2) for Male Puberty Timing. For Age at Menarche, full summary statistics were available [21] and used for reverse MR analyses and MR-APSS (see below). Full summary statistics for common EF were obtained from the GWAS Catalog (GCST90162547_buildGRCh37.tsv) [22].

All effects of the SNPs used as instrumental variables were harmonized between the exposure and outcome, ensuring that each effect corresponded to the same allele. Palindromic SNPs were assumed to be reported on the forward strand. Following harmonization, the inverse-variance weighted (IVW) effect estimate was derived by combining (in a manner analogous to fixed-effects meta-analysis) each SNP’s effect for the exposure on the outcome [23].

*Sensitivity analyses*

To address pleiotropy—which can violate MR assumptions—potential pleiotropic SNPs were identified by MR-PRESSO and excluded from all further analyses. The MR-PRESSO (version 1.0) approach identifies potential pleiotropic outliers by comparing the observed residual sum of squares with a simulated (Gaussian) distribution of residuals and excludes these outliers [24]. After exclusion of potential pleiotropic SNPs, the inverse-variance weighted (IVW) effect estimate was derived by combining (in a manner analogous to fixed-effects meta-analysis) each SNP’s effect for the exposure on the outcome [23].

To additional address potential pleiotropy, the following robust or pleiotropy-correcting methods were used as sensitivity analyses and calculated after exclusion of potential pleiotropic variants: MR-Egger, Penalized Weighted Median, and Lasso Penalization. MR-Egger addresses directional pleiotropy by incorporating an intercept in the regression model and thereby allows all variants to be invalid, but it is again less powerful and is sensitive to violations of the Instrument Strength Independent of Direct Effect (InSIDE) assumption, which requires an absence of correlation between a genetic variant’s causal effect on the outcome and its direct (pleiotropic) effect [25]. MR-Egger also relies on the no-measurement-error (NOME) assumption, which treats genetic variant–exposure associations as being measured without error. Violations of these assumptions can be explored using the *I²* statistic; values of *I²* < 0.9 indicate a greater than 10% relative bias in the MR-Egger estimate [26]. The Weighted Median method can yield valid estimates as long as the majority (or plurality, in the mode-based approach) of instruments that share a similar causal effect adhere to MR assumptions [27, 28]. As a modification of this approach, the Penalized Weighted Median method was calculated, were the weight of each SNP to derive the effect estimate is penalized based on heterogeneity statistics, which indicate potential pleiotropy [29]. Further, effect estimates under the Lasso Penalization method were calculated [30]. This method introduces an intercept term for each SNP to the IVW method which represents the direct association between the SNP and the outcome, which provides a violation of the *exclusion restriction* assumption. Weighted linear regression models are used to estimate this intercept term, which is subject to Lasso penalization [30]. This penalization encourages sparsity by shrinking the intercept toward zero. SNPs with a intercept term equal to zero are identified as valid instruments and only these are used to calculate the IVW effect estimate [30]. Harmonization procedures and the calculation of IVW, MR-Egger, Weighted Median, and Penalized Weighted Median causal effect estimates were calculated using the TwoSampleMR package (version 0.6.8) [31]. Calculation of the *I²* statistic and of the Lasso Penalization method was performed using the MendelianRandomization package (version 0.10.0) [32].

As an additional sensitivity analysis, the MR-APSS method (version 0.0.0.9) was applied [33]. This method uses a foreground-background model approach, with the background model accounting for correlated pleiotropy and sample structure, while the foreground model is used to estimate the causal effect and correct for uncorrelated pleiotropy [33]. By employing linkage disequilibrium (LD) score regression (LDSC) as a background model, MR-APSS separates pleiotropic effects (which are associated with LD) from population structure effects (which do not correlate with LD) and thus is able to account for sample overlap [33]. Recent comparisons indicate that MR-APSS often performs better in terms of replicability, estimate accuracy, and type I error control than other MR approaches [34]. For the foreground model, MR-APSS uses its own definition of valid genetic instruments with a more relaxed significance threshold (p < 5×10^-5^ by default) and applies LD pruning (r² < 0.001 by default) for near-independence of variants [33]. Therefore, no prior exclusion of potential pleiotropic SNPs wit MR-PRESSO was performed.

*Multivariable MR analysis with BMI*

Multivariable Mendelian randomization (MVMR) analyses were undertaken to address BMI-driven pleiotropy. For this purpose, sex-specific GWAS data on BMI were obtained from 434,794 female and 374,756 male participants from 29 cohorts within the Genetic Investigation of ANthropometric Traits (GIANT) consortium and the UK Biobank [35]. After harmonizing these data using the TwoSampleMR package, and after exclusion of potential pleiotropic SNPs with MR-PRESSO [24]. MVMR estimates were derived for inverse-variance weighted (IVW), MR-Egger, Weighted Median, and Lasso Penalization approaches, employing the MendelianRandomization package (version 0.10.0) ) [32].

*Quality control measures*

When full summary statistics were accessible, the CheckSumStats package (version 0.0.0.9) [36] was employed to verify correct annotation of the effect allele and to confirm that the data originated from European-ancestry populations. Scatter plots, depicting each SNP’s effect on male puberty timing relative to its effect on the outcome of interest, and funnel plots, showing MR estimates (per SNP) against the inverse of their standard errors, were generated (using the TwoSampleMR package) to visually check for possible assumption violations or single-variant distortions [37]. Results given by the TwoSampleMR package were cross-checked using the MendelianRandomization package. Scatter plots, depicting each SNP’s effect on male puberty timing relative to its effect on the outcome of interest, and funnel plots, showing MR estimates (per SNP) against the inverse of their standard errors, were generated to visually check for possible assumption violations or single-variant distortions.

*Instrumental variables*

The proportion of the variance (PVE) explained by the SNPs used as instrumental variables for each exposure were calculated with the formula provided by Shim, et al [38]. Of 1,080 independent lead SNPs identified for Age at Menarche, 971 were genome-wide significant (p < 5×10^−8) in the European-only subcohort [19]. These explained PVE=10.7% of the variance for age at menarche. Of these 971 SNPs, 832 SNPs (85.7%) with a PVE=9.4% were available in the outcome GWAS. Given the minimal reduction in explained variance and potential for introducing linkage error or heterogeneity, we opted not to include proxy SNPs. The MR-PRESSO global test for pleiotropy was significant (RSSobs=2609.5, p<1x10^-4^). MR-PRESSO identified 27 potential pleiotropic SNPs, which were excluded from further analysis, resulting in 805 SNPs serving as the instrumental variable for age at menarche.

For male puberty timing, 76 genome-wide significant independent lead SNPs with a total PVE of 2.2% were identified by the GWAS of Hollis, et al. [20]. Of these, 72 (94.7%, PVE 2.0%) were available in the outcome GWAS. The MR-PRESSO global test for pleiotropy was significant (RSSobs=199.9, p<2x10^-4^), and MR-PRESSO identified one potential pleiotropic SNP, which was excluded from further analysis, resulting in 71 SNPs serving as the instrumental variable for male puberty timing.

### Supplementary Results

**Supplementary Results 2 – Results of Mendelian Randomization analyses**

*Age at menarche*

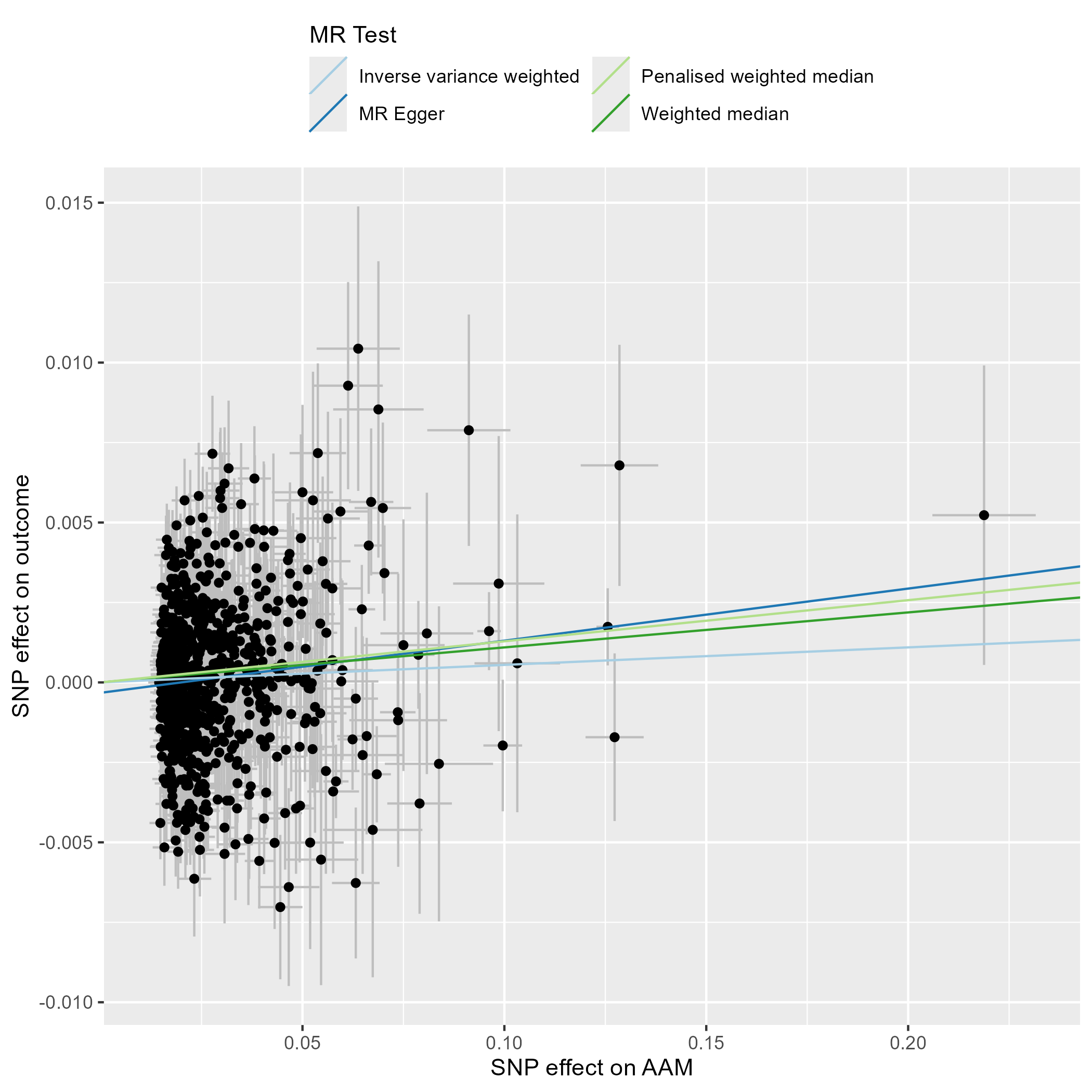

**Supplementary Figure S14 – Scatter Plot – Age at Menarche.** This scatter plot depicts the effect estimates (beta, error bars represent the standard error) of each genetic variant included in the analysis on the exposure (Age at Menarche [19]) and the outcome (common Executive Function factor, [39])

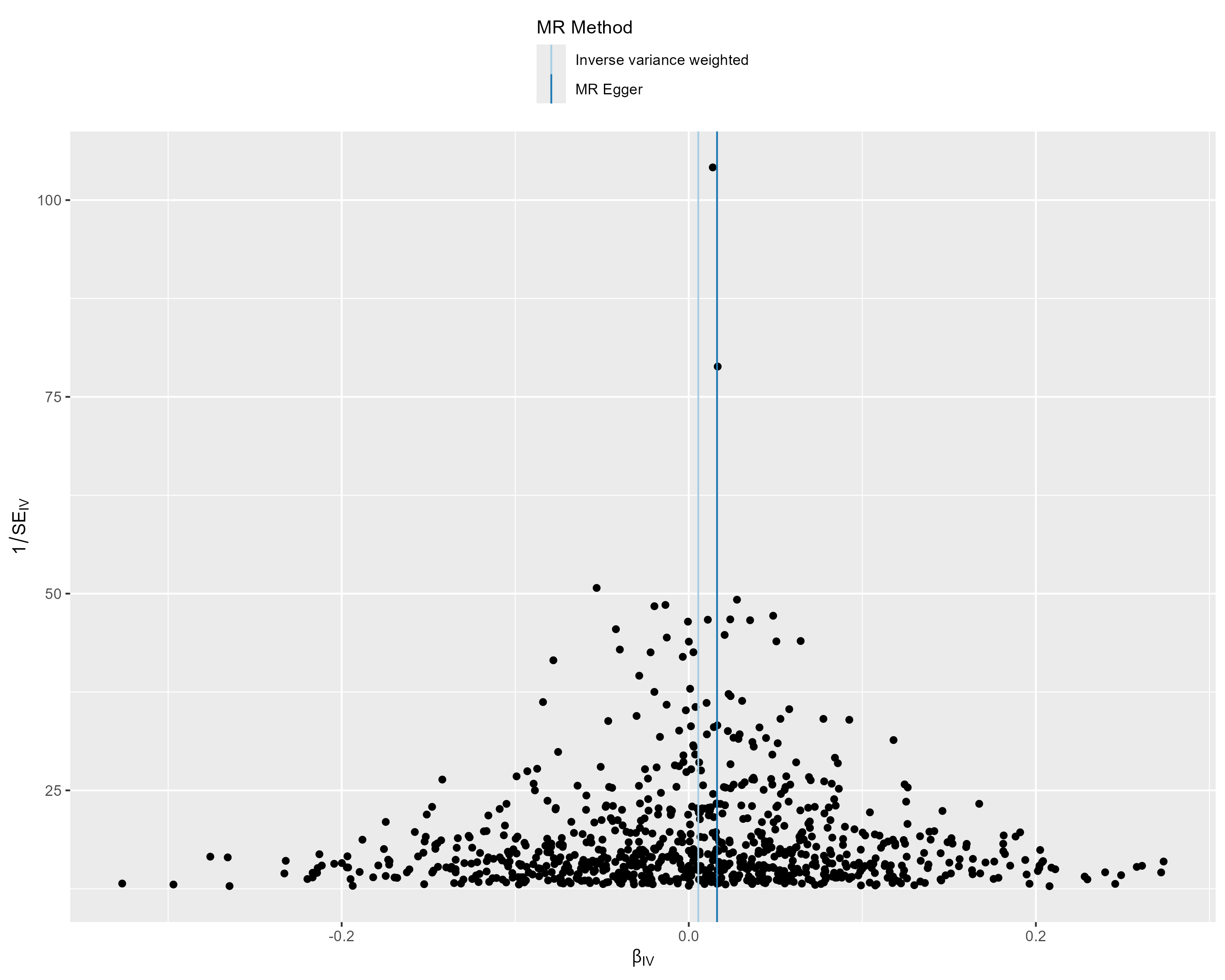

**Supplementary Figure S15 – Funnel Plot – Age at Menarche.** This funnel plot illustrates the precision of each genetic variant (as measured by the inverse of the standard error (SEIV)) and their MR effect estimate (βIV).

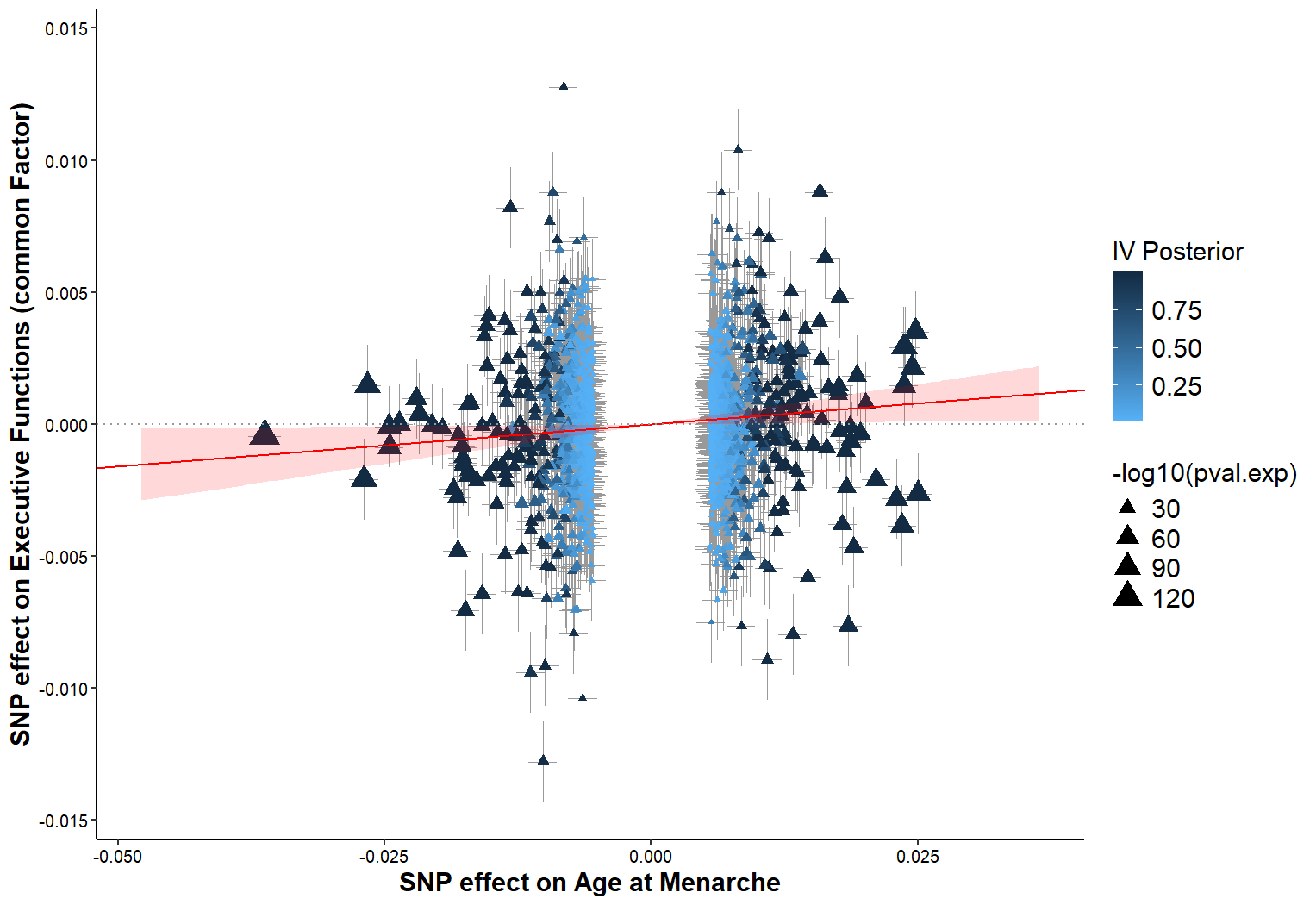
**Supplementary Figure S16 – MR-APSS Results.** Triangles represent SNP effects on Age at Menarche (exposure) and EF (outcome). Red line: estimated causal effect (95% CI shaded).MR-APSS uses a background model to account for correlated pleiotropy and sample structure (including sample overlap, cryptic relatedness, and sample stratification) and a foreground model to estimate the causal effect and correct for uncorrelated pleiotropy. The posterior probability (IV Posterior) quantifies the likelihood that a genetic variant (SNP) belongs to the foreground model (valid IV with true causal signal, represented in dark blue or black) versus the background model (represented in light blue). MR-APSS identified 1,447 independent ((r² < 0.001) genetic variants strongly related to the exposure (p < 5×10^-5^), where 413.6 SNPs contributed to the foreground model. MR-APSS estimated a causal effect of Age at Menarche on EF performance, with later Age at Menarche leading to higher EF performance ((b)=0.032 per one-year increase in Age at Menarche, 95%-CI [0.003, 0.060]).

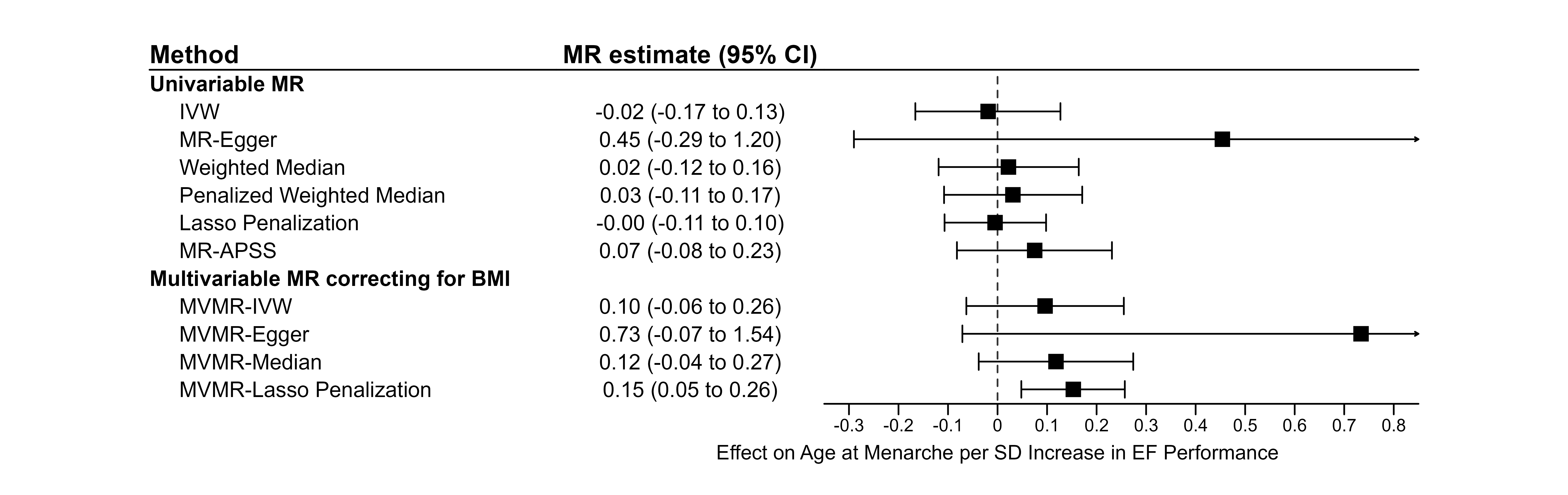
 **Supplementary Figure S17 – Reverse Mendelian Randomization analysis.** To analyze whether EF influences puberty timing, Mendelian Randomization analysis with the common EF factor as the exposure and Age at Menarche as the outcome was conducted. The instrumental variable for ‘Executive Function’ comprised by the 129 independent, genome-wide significant lead SNPs as identified by Hatoum, et al. [39]. Univariable Mendelian randomization (MR) analyses did not provide evidence for a causal effect of EF on age at menarche. In multivariable MR (MVMR) analyses accounting for BMI-related pleiotropy, results were inconsistent. Notably, only the Lasso Penalization method yielded a significant effect estimate suggestive of a potential effect of EF on age at menarche.

*Male Puberty Timing*

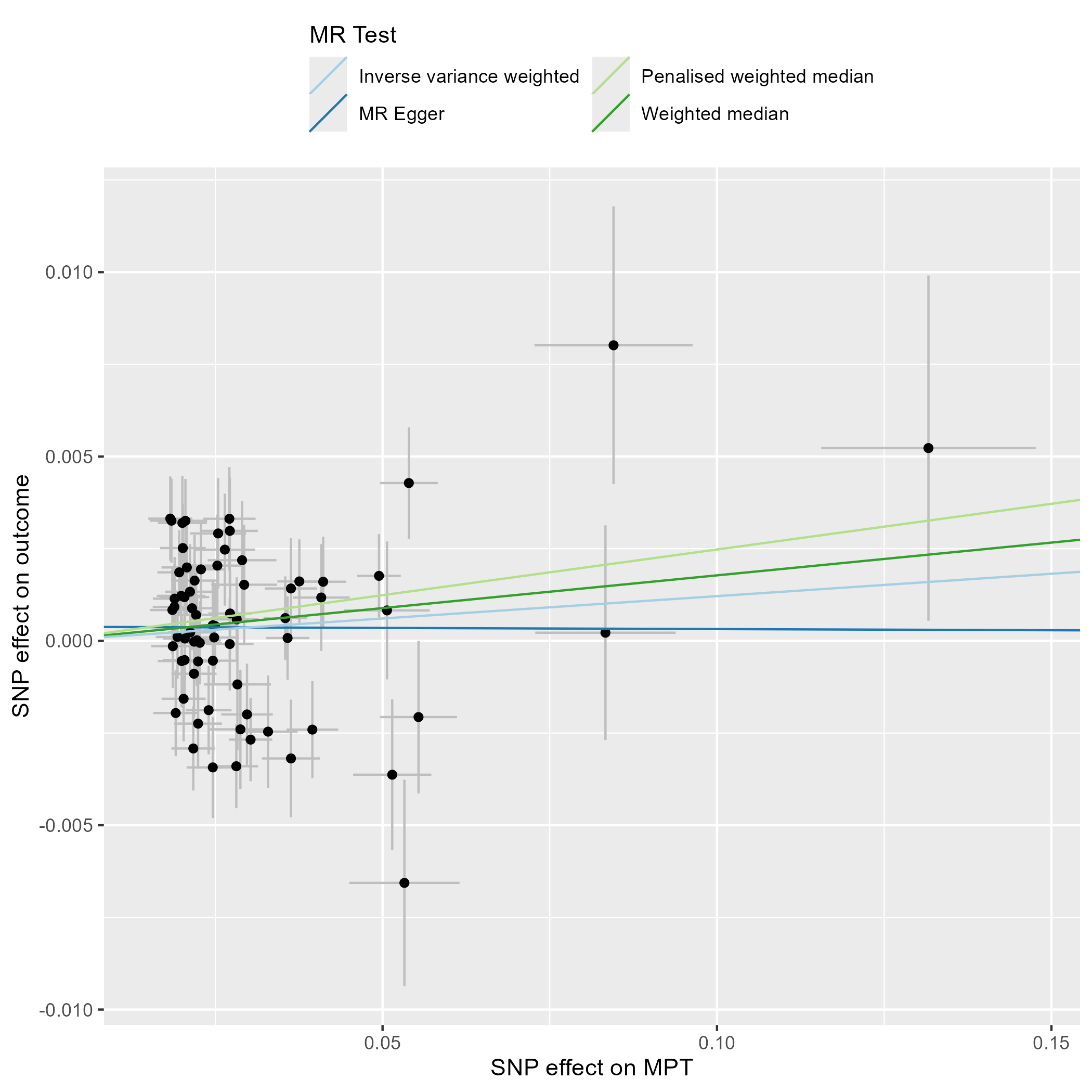

**Supplementary Figure S18 – Scatter Plot – Male Puberty Timing.** This scatter plot depicts the effect estimates (beta, error bars represent the standard error) of each genetic variant included in the analysis on the exposure (Male Puberty Timing [20]) and the outcome (common Executive Function factor, [39])

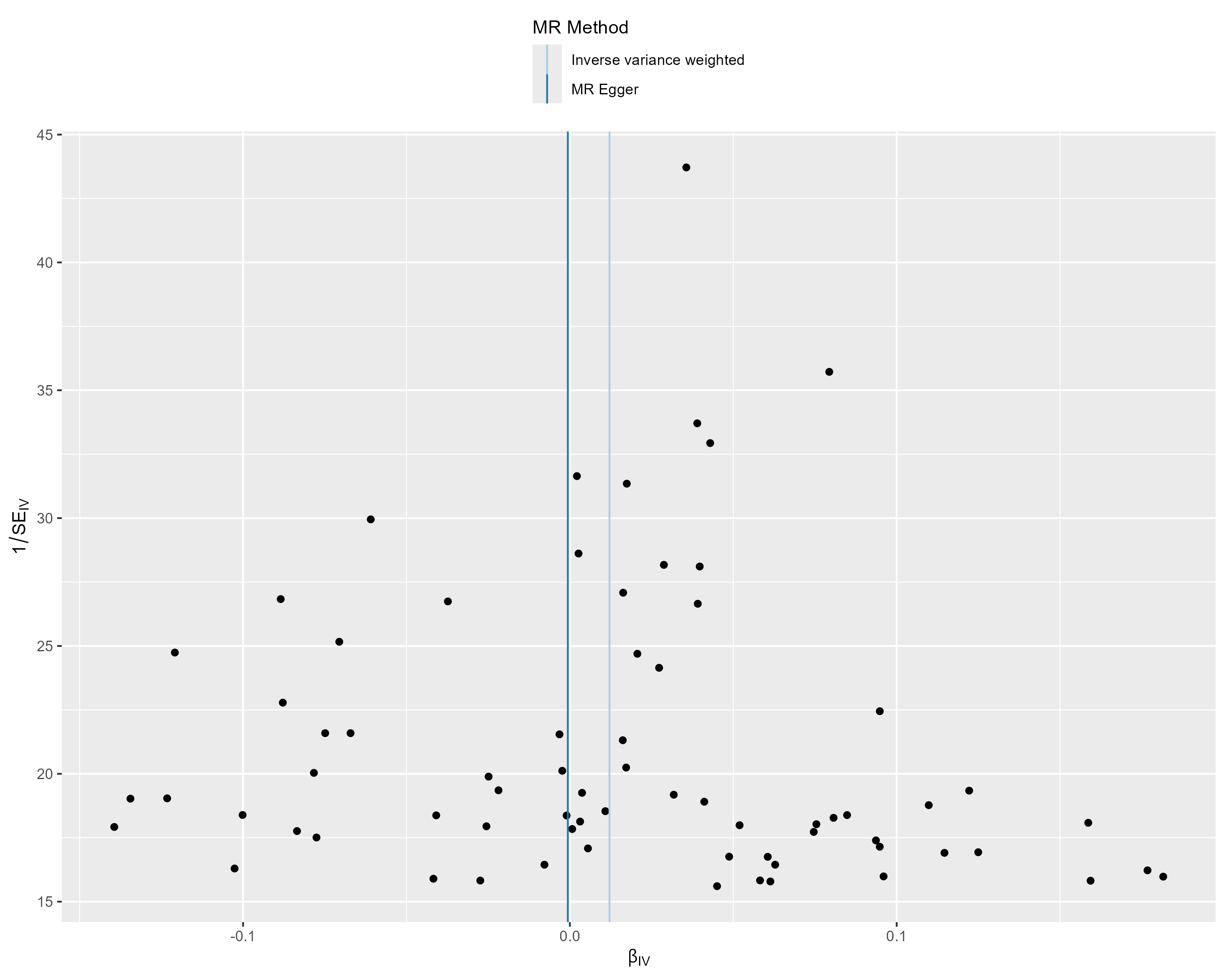

**Supplementary Figure S19 – Funnel Plot – Male Puberty Timing.** This funnel plot illustrates the precision of each genetic variant (as measured by the inverse of the standard error (SEIV)) and their MR effect estimate (βIV).

**Supplementary References**

1.	Petersen AC, Crockett L, Richards M, Boxer A (1988) A self-report measure of pubertal status: Reliability, validity, and initial norms. Journal of youth and adolescence 17:117-133

2.	Textor J, Hardt J, Knüppel S (2011) DAGitty: a graphical tool for analyzing causal diagrams. Epidemiology 22:745

3.	Lipsky AM, Greenland S (2022) Causal directed acyclic graphs. Jama 327:1083-1084

4.	Assari S, Boyce S, Bazargan M, Caldwell CH (2020) Race, socioeconomic status, and sex hormones among male and female American adolescents. Reproductive Medicine 1:8

5.	Reinehr T, Roth CL (2019) Is there a causal relationship between obesity and puberty? The Lancet Child & Adolescent Health 3:44-54

6.	Finn AS, Minas JE, Leonard JA, Mackey AP, Salvatore J, Goetz C, et al. (2017) Functional brain organization of working memory in adolescents varies in relation to family income and academic achievement. Developmental Science 20:e12450

7.	Decker AL, Duncan K, Finn AS, Mabbott DJ (2020) Children’s family income is associated with cognitive function and volume of anterior not posterior hippocampus. Nature Communications 11:4040

8.	Miller S, DeBoer M, Scharf R (2018) Executive functioning in low birth weight children entering kindergarten. Journal of perinatology 38:98-103

9.	Peng Q, Qiu W, Li Z, Zhao J, Zhu C (2024) Fetal genetically determined birth weight plays a causal role in earlier puberty timing: evidence from human genetic studies. Human Reproduction 39:792-800

10.	Likhitweerawong N, Louthrenoo O, Boonchooduang N, Tangwijitsakul H, Srisurapanont M (2022) Bidirectional prediction between weight status and executive function in children and adolescents: a systematic review and meta‐analysis of longitudinal studies. Obesity Reviews 23:e13458

11.	Ogden CL, Kuczmarski RJ, Flegal KM, Mei Z, Guo S, Wei R, et al. (2002) Centers for Disease Control and Prevention 2000 growth charts for the United States: improvements to the 1977 National Center for Health Statistics version. Pediatrics 109:45-60

12.	Van Buuren S, Groothuis-Oudshoorn K (2011) mice: Multivariate imputation by chained equations in R. Journal of statistical software 45:1-67

13.	Marcoulides KM, Raykov T (2019) Evaluation of variance inflation factors in regression models using latent variable modeling methods. Educational and psychological measurement 79:874-882

14.	Burnham KP, Anderson DR (2004) Multimodel inference: understanding AIC and BIC in model selection. Sociological methods & research 33:261-304

15.	Lewis F, Butler A, Gilbert L (2011) A unified approach to model selection using the likelihood ratio test. Methods in ecology and evolution 2:155-162

16.	Bates D, Mächler M, Bolker B, Walker S (2015) Fitting linear mixed-effects models using lme4. Journal of statistical software 67:1-48

17.	Kuznetsova A, Brockhoff PB, Christensen RH (2017) lmerTest package: tests in linear mixed effects models. Journal of statistical software 82:1-26

18.	Fox J, Weisberg S (2018) An R companion to applied regression. Sage publications

19.	Kentistou KA, Kaisinger LR, Stankovic S, Vaudel M, Mendes de Oliveira E, Messina A, et al. (2024) Understanding the genetic complexity of puberty timing across the allele frequency spectrum. Nature genetics 56:1397-1411

20.	Hollis B, Day FR, Busch AS, Thompson DJ, Soares ALG, Timmers PR, et al. (2020) Genomic analysis of male puberty timing highlights shared genetic basis with hair colour and lifespan. Nature communications 11:1536

21.	Kentistou K, Day F, Perry J, Ong K (2024) Research data supporting:" Understanding the genetic complexity of puberty timing across the allele frequency spectrum".

22.	MacArthur J, Bowler E, Cerezo M, Gil L, Hall P, Hastings E, et al. (2017) The new NHGRI-EBI Catalog of published genome-wide association studies (GWAS Catalog). Nucleic acids research 45:D896-D901

23.	Burgess S, Butterworth A, Thompson SG (2013) Mendelian randomization analysis with multiple genetic variants using summarized data. Genetic epidemiology 37:658-665

24.	Verbanck M, Chen C-Y, Neale B, Do R (2018) Detection of widespread horizontal pleiotropy in causal relationships inferred from Mendelian randomization between complex traits and diseases. Nature genetics 50:693-698

25.	Bowden J, Davey Smith G, Burgess S (2015) Mendelian randomization with invalid instruments: effect estimation and bias detection through Egger regression. International journal of epidemiology 44:512-525

26.	Bowden J, Del Greco M F, Minelli C, Davey Smith G, Sheehan NA, Thompson JR (2016) Assessing the suitability of summary data for two-sample Mendelian randomization analyses using MR-Egger regression: the role of the I 2 statistic. International journal of epidemiology 45:1961-1974

27.	Bowden J, Davey Smith G, Haycock PC, Burgess S (2016) Consistent estimation in Mendelian randomization with some invalid instruments using a weighted median estimator. Genetic epidemiology 40:304-314

28.	Hartwig FP, Davey Smith G, Bowden J (2017) Robust inference in summary data Mendelian randomization via the zero modal pleiotropy assumption. International journal of epidemiology 46:1985-1998

29.	Burgess S, Bowden J, Dudbridge F, Thompson SG (2016) Robust instrumental variable methods using multiple candidate instruments with application to Mendelian randomization. arXiv preprint arXiv:1606.03729

30.	Rees JM, Wood AM, Dudbridge F, Burgess S (2019) Robust methods in Mendelian randomization via penalization of heterogeneous causal estimates. PloS one 14:e0222362

31.	Hemani G, Zheng J, Elsworth B, Wade KH, Haberland V, Baird D, et al. (2018) The MR-Base platform supports systematic causal inference across the human phenome. elife 7:e34408

32.	Yavorska OO, Burgess S (2017) MendelianRandomization: an R package for performing Mendelian randomization analyses using summarized data. International journal of epidemiology 46:1734-1739

33.	Hu X, Zhao J, Lin Z, Wang Y, Peng H, Zhao H, et al. (2022) Mendelian randomization for causal inference accounting for pleiotropy and sample structure using genome-wide summary statistics. Proceedings of the National Academy of Sciences 119:e2106858119

34.	Hu X, Cai M, Xiao J, Wan X, Wang Z, Zhao H, et al. (2024) Benchmarking Mendelian randomization methods for causal inference using genome-wide association study summary statistics. The American Journal of Human Genetics 111:1717-1735

35.	Pulit SL, Stoneman C, Morris AP, Wood AR, Glastonbury CA, Tyrrell J, et al. (2019) Meta-analysis of genome-wide association studies for body fat distribution in 694 649 individuals of European ancestry. Human molecular genetics 28:166-174

36.	Haycock PC, Borges MC, Burrows K, Lemaitre RN, Harrison S, Burgess S, et al. (2023) Design and quality control of large-scale two-sample Mendelian randomization studies. International journal of epidemiology 52:1498-1521

37.	Burgess S, Bowden J, Fall T, Ingelsson E, Thompson SG (2017) Sensitivity analyses for robust causal inference from Mendelian randomization analyses with multiple genetic variants. Epidemiology (Cambridge, Mass.) 28:30

38.	Shim H, Chasman DI, Smith JD, Mora S, Ridker PM, Nickerson DA, et al. (2015) A multivariate genome-wide association analysis of 10 LDL subfractions, and their response to statin treatment, in 1868 Caucasians. PLoS one 10:e0120758

39.	Hatoum AS, Morrison CL, Mitchell EC, Lam M, Benca-Bachman CE, Reineberg AE, et al. (2023) Genome-wide association study shows that executive functioning is influenced by GABAergic processes and is a neurocognitive genetic correlate of psychiatric disorders. Biological psychiatry 93:59-70
