## Supplemental Tables S8 and S9 for "Puberty Timing and Cognitive Functioning: Insights from the Adolescent Brain Cognitive Development (ABCD) Study and Mendelian Randomization"

| **Supplementary Table S8.** Comparison of Effect Estimates for Effect of Puberty Timing on Cognition across all Timepoints (‘Overall’) per Model Adjustments. | | | | | | | | | | | |
| --- | --- | --- | --- | --- | --- | --- | --- | --- | --- | --- | --- |
|  |  | Crude model | | | Adjusted model | | | | Adjusted + CBCL_int/ext_ model | | |
|  | N_obs_ | β | 95%-CI | p_raw_ | β | 95%-CI | p_raw_ | p_FDRcorr_ | β | 95%-CI | p_raw_ |
| **Females (N = 4,879)** |  |  |  |  |  |  |  |  |  |  |  |
| NIH Toolbox® Flanker Task | 9,606 | -0.10 | (-0.13, -0.08) | < 0.001 | -0.02 | (-0.05, 0.00) | 0.071 | 0.098 | -0.02 | (-0.05, 0.00) | 0.094 |
| NIH Toolbox® Pattern Comparison | 9,588 | -0.06 | (-0.08, -0.04) | < 0.001 | -0.00 | (-0.02, 0.02) | 0.987 | 0.987 | 0.00 | (-0.02, 0.02) | 0.961 |
| NIH Toolbox® Picture Sequence | 11,020 | -0.14 | (-0.16, -0.11) | < 0.001 | **-0.04** | **(-0.06, -0.01)** | **0.003** | **0.015** | -0.03 | (-0.06, -0.01) | 0.008 |
| NIH Toolbox® List Sorting | 6,756 | -0.15 | (-0.17, -0.12) | < 0.001 | **-0.03** | **(-0.06, -0.01)** | **0.015** | **0.030** | -0.03 | (-0.06, -0.00) | 0.022 |
| RAVLT – Acquisition | 9,147 | -0.14 | (-0.17, -0.12) | < 0.001 | **-0.05** | **(-0.07, -0.02)** | **0.001** | **0.011** | -0.04 | (-0.07, -0.02) | 0.001 |
| RAVLT – Delayed Recall | 9,104 | -0.14 | (-0.16, -0.11) | < 0.001 | **-0.03** | **(-0.06, -0.01)** | **0.017** | **0.030** | -0.03 | (-0.06, -0.00) | 0.022 |
| RAVLT – Retroactive Interference | 9,123 | 0.03 | (0.01, 0.05) | 0.015 | -0.00 | (-0.03, 0.02) | 0.819 | 0.947 | -0.00 | (-0.03, 0.02) | 0.914 |
| LMT – Accuracy | 11,039 | -0.13 | (-0.15, -0.11) | < 0.001 | **-0.03** | **(-0.05, -0.01)** | **0.015** | **0.030** | -0.03 | (-0.05, -0.00) | 0.029 |
| LMT – Response Time | 11,035 | -0.03 | (-0.05, -0.01) | 0.001 | **-0.03** | **(-0.05, -0.01)** | **0.004** | **0.015** | -0.03 | (-0.05, -0.01) | 0.005 |
| **Males (N = 5,295)** |  |  |  |  |  |  |  |  |  |  |  |
| NIH Toolbox® Flanker Task | 10,533 | -0.08 | (-0.11, -0.06) | < 0.001 | -0.02 | (-0.04, 0.00) | 0.058 | 0.087 | -0.02 | (-0.04, 0.00) | 0.071 |
| NIH Toolbox® Pattern Comparison | 10,513 | -0.04 | (-0.06, -0.02) | < 0.001 | -0.00 | (-0.02, 0.02) | 0.840 | 0.945 | -0.00 | (-0.02, 0.02) | 0.898 |
| NIH Toolbox® Picture Sequence | 12,051 | -0.10 | (-0.12, -0.08) | < 0.001 | **-0.03** | **(-0.05, -0.01)** | **0.014** | **0.030** | -0.03 | (-0.05, -0.00) | 0.023 |
| NIH Toolbox® List Sorting | 7,409 | -0.13 | (-0.15, -0.10) | < 0.001 | **-0.04** | **(-0.06, -0.01)** | **0.004** | **0.015** | -0.03 | (-0.06, -0.01) | 0.006 |
| RAVLT – Acquisition | 9,922 | -0.12 | (-0.15, -0.10) | < 0.001 | **-0.04** | **(-0.06, -0.01)** | **0.003** | **0.015** | -0.04 | (-0.06, -0.01) | 0.004 |
| RAVLT – Delayed Recall | 9,864 | -0.12 | (-0.14, -0.09) | < 0.001 | **-0.03** | **(-0.06, -0.01)** | **0.007** | **0.022** | -0.03 | (-0.06, -0.01) | 0.010 |
| RAVLT – Retroactive Interference | 9,880 | 0.01 | (-0.01, 0.03) | 0.443 | -0.01 | (-0.03, 0.01) | 0.511 | 0.657 | -0.01 | (-0.03, 0.01) | 0.439 |
| LMT – Accuracy | 12,060 | -0.10 | (-0.12, -0.08) | < 0.001 | -0.02 | (-0.04, -0.00) | 0.048 | 0.079 | -0.02 | (-0.04, 0.00) | 0.068 |
| LMT – Response Time | 12,056 | -0.01 | (-0.03, 0.01) | 0.218 | -0.00 | (-0.02, 0.02) | 0.959 | 0.987 | 0.00 | (-0.02, 0.02) | 0.943 |

This table presents the association between Puberty Timing—measured as standardized residuals from regressing the Pubertal Development Scale (PDS) total score (at Baseline) on age, indicating pubertal maturation relative to same-age, same-sex peers—and cognitive task performance. Shown are standardized effect sizes (SD change in cognitive performance per SD increase in Puberty Timing), estimated using mixed linear models across all timepoints (Baseline, 2-Year, and 4-Year Follow-Ups). The ‘Crude Model’ only included random intercepts for subject, timepoint, and family nested within site. The ‘Adjusted Model’ additionally accounted for BMI-SDS, race/ethnicity, family income, and birthweight as fixed effects. The ‘Adjusted + CBCLint/ext Model’ further included raw CBCL internalizing and externalizing scores. For the primary outcome (Puberty Timing effect on cognitive outcomes in the Adjusted Model), FDR-corrected p-values (p_FDRcorr_) were computed and values < 0.05 were considered statistically significant and are shown in bold. BMI-SDS = BMI-Standard Deviation Score; N_obs_ = Number of Observations; 95%-CI = 95% Confidence Interval. p_raw_ = p-values without FDR-correction.

| **Supplementary Table S9.** Comparison of Effect Estimates for Effect of Puberty Timing on Cognitive Tasks per Timepoint. | | | | | | | | | | | | |
| --- | --- | --- | --- | --- | --- | --- | --- | --- | --- | --- | --- | --- |
|  | Baseline | | | | 2-Years Follow-Up | | | | 4-Years Follow-Up | | | |
|  | N_obs_ | β | 95%-CI | p | N_obs_ | β | 95%-CI | p | N_obs_ | β | 95%-CI | p |
| **Females (N = 4,879)** |  |  |  |  |  |  |  |  |  |  |  |  |
| NIH Toolbox® Flanker Task | 4,879 | -0.03 | (-0.06, 0.01) | 0.110 | 3,363 | -0.02 | (-0.06, 0.01) | 0.220 | 1,364 | 0.01 | (-0.05, 0.07) | 0.782 |
| NIH Toolbox® Pattern Comparison | 4,879 | -0.01 | (-0.04, 0.03) | 0.665 | 3,351 | 0.01 | (-0.03, 0.05) | 0.508 | 1,358 | 0.01 | (-0.05, 0.08) | 0.678 |
| NIH Toolbox® Picture Sequence | 4,879 | -0.02 | (-0.05, 0.02) | 0.310 | 4,289 | -0.06 | (-0.09, -0.03) | < 0.001 | 1,852 | -0.04 | (-0.09, 0.02) | 0.196 |
| NIH Toolbox® List Sorting | 4,879 | -0.04 | (-0.07, -0.01) | 0.015 | **-** | **-** | **-** | **-** | 1,845 | -0.02 | (-0.07, 0.03) | 0.480 |
| RAVLT – Acquisition | 4,879 | -0.04 | (-0.08, -0.01) | 0.007 | 4,268 | -0.06 | (-0.09, -0.02) | 0.001 | **-** | **-** | **-** | **-** |
| RAVLT – Delayed Recall | 4,879 | -0.04 | (-0.07, -0.01) | 0.015 | 4,225 | -0.03 | (-0.06, 0.01) | 0.119 | **-** | **-** | **-** | **-** |
| RAVLT – Retroactive Interference | 4,879 | -0.00 | (-0.04, 0.03) | 0.796 | 4,244 | -0.00 | (-0.04, 0.03) | 0.914 | **-** | **-** | **-** | **-** |
| LMT – Accuracy | 4,879 | -0.03 | (-0.06, 0.01) | 0.102 | 4,315 | -0.03 | (-0.06, 0.00) | 0.072 | 1,845 | -0.08 | (-0.14, -0.03) | 0.002 |
| LMT – Response Time | 4,879 | -0.04 | (-0.07, -0.01) | 0.019 | 4,314 | -0.02 | (-0.06, 0.01) | 0.181 | 1,842 | -0.07 | (-0.13, -0.02) | 0.007 |
| **Males (N = 5,295)** |  |  |  |  |  |  |  |  |  |  |  |  |
| NIH Toolbox® Flanker Task | 5,295 | -0.02 | (-0.04, 0.01) | 0.273 | 3,745 | -0.03 | (-0.07, 0.00) | 0.086 | 1,493 | -0.02 | (-0.08, 0.04) | 0.542 |
| NIH Toolbox® Pattern Comparison | 5,295 | 0.00 | (-0.03, 0.03) | 0.799 | 3,723 | -0.02 | (-0.06, 0.02) | 0.296 | 1,495 | 0.03 | (-0.03, 0.09) | 0.353 |
| NIH Toolbox® Picture Sequence | 5,295 | -0.02 | (-0.05, 0.01) | 0.110 | 4,684 | -0.03 | (-0.06, 0.00) | 0.080 | 2,072 | -0.05 | (-0.10, -0.00) | 0.042 |
| NIH Toolbox® List Sorting | 5,295 | -0.04 | (-0.07, -0.01) | 0.007 | **-** | - | - | - | 2,077 | -0.04 | (-0.09, 0.01) | 0.112 |
| RAVLT – Acquisition | 5,295 | -0.04 | (-0.07, -0.01) | 0.006 | 4,627 | -0.03 | (-0.06, 0.00) | 0.053 | - | - | - | - |
| RAVLT – Delayed Recall | 5,295 | -0.04 | (-0.07, -0.01) | 0.006 | 4,569 | -0.02 | (-0.05, 0.01) | 0.155 | - | - | - | - |
| RAVLT – Retroactive Interference | 5,295 | -0.00 | (-0.03, 0.03) | 0.990 | 4,585 | -0.02 | (-0.05, 0.02) | 0.312 | - | - | - | - |
| LMT – Accuracy | 5,295 | -0.00 | (-0.03, 0.02) | 0.785 | 4,728 | -0.03 | (-0.06, -0.00) | 0.046 | 2,037 | -0.06 | (-0.11, -0.01) | 0.011 |
| LMT – Response Time | 5,295 | 0.00 | (-0.03, 0.03) | 0.837 | 4,727 | 0.01 | (-0.02, 0.04) | 0.550 | 2,034 | -0.03 | (-0.08, 0.02) | 0.270 |

This table presents the association between Puberty Timing (at Baseline) and cognitive task performance per timepoint. Shown are standardized effect sizes (SD change in cognitive performance per SD increase in Puberty Timing), estimated using mixed linear models across all timepoints (Baseline, 2-Year, and 4-Year Follow-Ups). All models were adjusted using BMI-SDS, race/ethnicity, family income, and birthweight as fixed effects and included random intercept for family nested within side. Please note that for the NIH Toolbox® List Sorting 2-Years Follow-Ups and for the RAVLT, 4-Years Follow-Ups were not available. BMI-SDS = BMI-Standard Deviation Score; Nobs = Number of Observations; 95%-CI = 95% Confidence Interval.
